# Estimating the strength of selection for new SARS-CoV-2 variants

**DOI:** 10.1101/2021.03.29.21254233

**Authors:** Christiaan H. van Dorp, Emma E. Goldberg, Nick Hengartner, Ruian Ke, Ethan O. Romero-Severson

**Author notes:** These authors contributed equally to this work (Christiaan H. van Dorp, Emma E. Goldberg).

## Abstract

Controlling the SARS-CoV-2 pandemic becomes increasingly challenging as the virus adapts to human hosts through the continual emergence of more transmissible variants. Simply observing that a variant is increasing in frequency is relatively straightforward, but more sophisticated methodology is needed to determine whether a new variant is a global threat and the magnitude of its selective advantage. We present three methods for quantifying the strength of selection for new and emerging variants of SARS-CoV-2 relative to the background of contemporaneous variants. These methods range from a detailed model of dynamics within one country to a broad analysis across all countries, and they include alternative explanations such as migration and drift. We find evidence for strong selection favoring the D614G spike mutation and B.1.1.7 (Alpha), weaker selection favoring B.1.351 (Beta), and no advantage of R.1 after it spreads beyond Japan. Cutting back data to earlier time horizons reveals large uncertainty very soon after emergence, but that estimates of selection stabilize after several weeks. Our results also show substantial heterogeneity among countries, demonstrating the need for a truly global perspective on the molecular epidemiology of SARS-CoV-2.

## Introduction

Recently, several genetic variants of SARS-CoV-2 have been identified that are either suspected or confirmed to have mutations that increase the contagiousness of the virus above the current circulating variants [1, 2, 3, 4]. For a short while after the emergence of SARS-CoV-2, it was believed that the adaptive evolution of SARS-CoV-2 was limited, as evidence for purifying selection was found at most sites, with the clear exception of position 614 of the spike protein [5]. However, the emergence and rise of more complex variants such as B.1.1.7 (Alpha) [3] and B.1.617.2 (Delta) [6] have shifted this understanding. As SARS-CoV-2 continues to adapt to transmission among humans, we can expect to see further mutations that alter the phenotype of the circulating virus [7]. Likewise, the gradual roll-out of vaccination programs globally is changing the immunological landscape, possibly leading to the emergence of escape strains that are partially or fully resistant to existing vaccinations [8, 9, 10]. Although this effect may be slowed by greater and more equitable vaccination [11, 12], new variants will likely continue to emerge into the future. Identifying new genetic variants of concern as they are arising will thus continue to be important to guide vaccination and other public health strategies.

Molecular epidemiology comprises theory and software for analyzing pathogen genetic sequence data [1, 2, 3, 13]. These methods allow us to peer beyond what is provided by traditional epidemiological data such as case counts and death time-series, into the substructure of an epidemic by tracking the emergence and transmission of new genetic variants. Given the extensive population mixing at both local and global levels for SARS-CoV-2, the time between the emergence of a new variant in one country and its global dissemination is short. With such rapid spread, the ongoing fight against COVID-19 needs new, global tools focused on rapid modeling and assessment of the risk associated with new strains of SARS-CoV-2 to support global public health action.

Distinguishing which new variants truly pose a greater threat—as opposed to the many mutations that are not advantageous to the virus—is the first, qualitative step. Further quantifying the selective advantage of a variant provides greater insight into how, or how aggressively, to deploy interventions. Several groups have investigated the selective advantage of particular SARS-CoV-2 variants, both qualitatively and quantitatively. The global spread of the D614G variant was first described by Korber et al. [14]. Specifically for the UK, the selection coefficient for D614G has been estimated using phylogenetic and phylodynamic methods [3], although the estimates from the various methods are highly variable. The increased infectiousness of D614G has also been functionally explained in terms of ACE2 receptor binding [15]. The selection coefficient of the B.1.1.7 (Alpha) variant has been estimated for England using a highly detailed deterministic epidemic model [16]. Phylodynamic approaches have led to similar results [2]. More worryingly, B.1.1.7 (Alpha) is associated with increased mortality [17] (but see also [18]). It has been estimated that the contagiousness of the Delta variant is higher than the Alpha variant [4]. As of September 2021, Delta replaced Alpha and became the dominant variant in many countries. The ability of the Delta variant to cause more severe diseases in unvaccinated individuals and breakthrough infections in vaccinated individuals becomes a major concern [19, 20]. These changes to the SARS-CoV-2 phenotype embodied in D614G, Alpha, and Delta likely represent only a small fraction of the phenotypic variability in the broader population.

In this paper we develop three methods for analyzing global sequence data to estimate the selective advantage of SARS-CoV-2 genetic variants. We compare their consistency, strengths, and weaknesses while applying them to four variants present in many countries. As discussed above, D614G and B.1.1.7 (Alpha) emerged relatively early and received much attention as they rapidly spread globally. We also analyze B.1.351 (Beta), which was initially detected in South Africa in October 2020 [1], and rapidly spread in that country and several others before the dominance of the Delta variant. Finally, we consider the R.1 lineage which was first detected in Japan [21]. Although it initially rose in frequency rapidly in Japan, it did not achieve global spread. By applying our methods to these variants of concerns with different epidemiological patterns, we test the robustness of results to different modeling assumptions, and we assess our different approaches as molecular surveillance tools.

## Methods

We use three analysis techniques to study the change in frequency over time of a SARS-CoV-2 genetic variant: isotonic regression, a population genetics model, and a stochastic epidemiological model. These methods represent trade-offs in mechanistic detail and computational efficiency. The first takes a descriptive approach to the rise and fall of variant frequency based on rejecting a null hypothesis of limited or no change in frequency. The second incorporates the processes of selection and migration in the context of an idealized deterministic population. The third additionally includes stochastic effects and more explicit epidemiological processes. By comparing results from these three methods, we assess the robustness of our findings to the assumptions of each. In all models, we compare a focal variant with the pool of circulating background variants. The focal and background variants are labeled mt (for “mutant”) and wt (for “wild type”) respectively. Our analysis scripts can be downloaded from https://github.com/eeg-lanl/sarscov2-selection.

### Data

Death incidence data was taken from the COVID-19 Data Repository curated by the Center for Systems Science and Engineering (CSSE) at Johns Hopkins University [22], aggregated by week to reduce noise. Data for B.1.1.7, B.1.351, and R.1 were downloaded from the GISAID website [23, 24] on 20 May 2021, which includes Pango lineage designations. The D614G variant data were taken from the Los Alamos COVID-19 Viral Genome Analysis Pipeline [14, 25] on 17 June 2021. For each focal variant, the number of observed sequences with that variant in each country sampled each day were counted as the mutant, and the total number of other sampled sequences were counted as wild type.

To obtain our best estimates of the strength of selection, we analyzed the first several months of data for each variant. The data and date ranges are shown in Figs. 2–4 and Figs. S7–S10. In order to assess how early positive selection on a variant could be detected, we also analyzed data cut back to different time horizons. Each time horizon was defined by the day on which the number of sequenced cases (in GISAID) of the new variant exceeded a particular threshold for the population genetics model. The threshold numbers of cases and corresponding dates are show in Figs. 6 and 8. We apply our time horizons to the date at which a sample was taken, rather than the date at which it was recorded in a database. For the isotonic regression and population genetic methods, which analyze data from many countries simultaneously, we include any country in which all of the following criteria were met by the time horizon: at least 20 cases of mt, at least 20 cases of wt, and at least 14 days with any sequence data. For the stochastic epidemiological model we used fixed 14 day increments to define consecutive time horizons (see Figs. 3 and 4).

### Isotonic regression

The logic behind the isotonic regression method is that, if a variant is under selection strong enough to be worrying, then we should see a continual increase in its relative frequency. That is, for a variant under selection in a given country, we should be able to reject the hypothesis that it shows no increase with respect to its background.

Let us consider modeling the time series of pairs 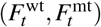 that count the number of samples identified as the new variant 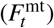 and all other background sequences 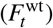 observed on a given day *t*. If we assume that the individuals whose SARS-CoV-2 virus is sequenced are randomly selected from the pool of infected individuals, then the number of observed variant sequences 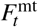, conditional on the total number of sequenced individuals 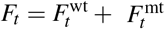, is binomial with probability *p*_*t*_ and sample size *F*_*t*_. If the mutations that define the new variant (i.e., the genotype) are neutral, being neither beneficial nor deleterious, then the proportion *p*_*t*_ performs a random walk with constant expectation. However, if the variant has an evolutionary advantage, then the proportion *p*_*t*_ will have an increasing expectation over time. Here, we use that observation to devise a statistical test for the null hypothesis that a genotype is not advantageous. This approach does not, however, provide us with an estimate of how advantageous a genotype is because it does not model the competition between variants.

Let (*t*_*i*_,*V*_*i*_), *i* = 1, …, *K* denote the date and variant *V*_*i*_ ∈ {wt, mt} from each of the *K* tested individuals. Our test is based on fitting isotonic logistic regressions to estimate a monotone non-decreasing probability *p*_*t*_ to that data, and using the logarithm of the likelihood ratio of the fitted isotonic model and model with constant *p*_*t*_ as the test statistic. Unlike in regular parametric cases, that statistic does not have an asymptotic chi-square distribution. For that reason, we empirically evaluate the null distribution of the test statistic by refitting the isotonic regression to 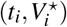, where 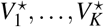 is a random permutation of the original data *V*_1_, …, *V*_*K*_. Fitting the isotonic logistic regression to *M* random permutations allows us to calculate empirically the country level p-value for the null hypothesis of no evolutionary advantage that are reported in Results. p-values were considered to be significant if they were below 0.05/*n* where *n* is the number of countries that met the inclusion criteria. These were calculated in R 3.6.3 using the package cgam [26] to perform the isotonic logistic regression.

### Population genetics model

The goal of this modeling approach is to provide a rapid means of estimating the selective advantage of a new genetic variant while also allowing that migration could provide an alternative explanation for its increase its frequency. We first describe the model within each country. Then we explain how we fit it in a hierarchical manner to data from multiple countries.

The model assumes that time is measured in discrete units of generations, which are non-overlapping. Within each generation, we let selection act first and then migration. Let *p* and *q* be the frequencies of new and background variants, respectively, at the beginning of the generation (*p* + *q* = 1). Then let *p*^*^ and *q*^*^ be the variant frequencies after selection, and *p*′ and *q*′ be the frequencies after migration and hence at the beginning of the next generation.

#### Selection

Define the absolute fitnesses of the two variants as *W*_wt_ = *β* and *W*_mt_ = *β* (1 + *s*). So *β* is the geometric growth rate in number of infected people for the original genotype, and *s* is the selective advantage (if *s* > 0) or disadvantage (if *s <* 0) of the new variant. Define *N*_mt_ = *Np* and *N*_wt_ = *Nq* as the numbers of infected people with each variant at the beginning of this generation, where *N* is the total number of infected people in the population. After the selective event, which is transmission of each of the variants to new hosts, the numbers of infected people become

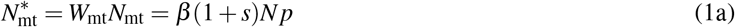

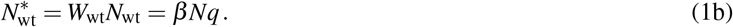

Even if transmission (and recovery) alters the number of infected people, this change in population size does not affect the new variant frequencies, i.e.,

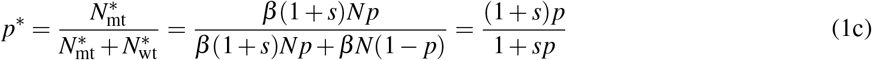

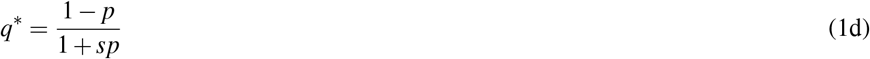

is independent of *N* and of *β* (under our assumption that generations are non-overlapping). So even with arbitrary changes in the number of infected people over time, this simple deterministic model can track only the variant frequencies. Of course, drift can have large effects when *N* is small, and also when a population of any size is growing rapidly. But we leave stochastic effects to our subsequent, more complex epidemic model. The selection-only version of our model, described thus far, is analogous to the logistic model fitting approach of Volz et al. [3], Chen et al. [27]. We proceed, however, to also incorporate migration and a hierarchical structure across countries, described next.

#### Migration

Next, a fraction *m* of our population is replaced by immigrants. That is, some number of infected people leave our population, and an equal number of infected people arrive from elsewhere. We say that immigration is balanced by emigration because we are applying this same model to many populations (countries) simultaneously, and travel itself does not change the total number of infected people.

The change in frequency of the new variant due to migration is

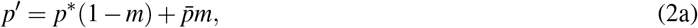

where 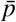 is the frequency of the new variant among the immigrants. To be most generous to the alternative explanation that immigration is the driving force behind increases in *p*, we set 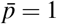 so

Note that if the number of infected people is increasing over time (*β* > 1 in the description of selection, above), our formulation with constant migration fraction *m* means that the number of infected travelers is also increasing over time.

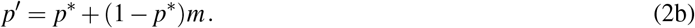

Putting together the total effects of selection and migration for this generation, by substituting Eq. (1c) into Eq. (2b),

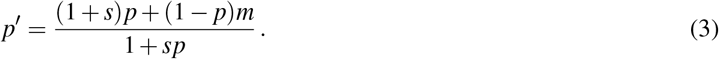

At any time *t*,

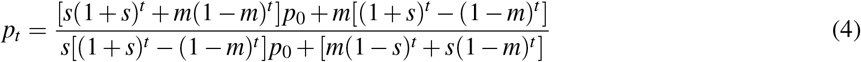

(see Eq. (A-6)). We define *t* = 0 as the time at which the new variant first appears in any country. Notice that without migration, *m* = 0, Eq. (4) reduces to the logistic model derived in [27].

#### Fitting to data

For each country, the data we use are the numbers of observations of the background 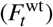 and the new variant 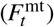 each day (*t*). We fit these data with Bayesian binomial regression, using Eq. (4), with country as a random effect. This yields separate estimates of *s, m*, and *p*_0_ for each country. When selection truly favors a variant due to its genetic composition, it should have a similar advantage in any country. There may be differences from country to country, though, due to chance effects. For example, if the early-infected people in one country happen to be from a demographic with higher transmission or a city with looser enforcement of social distancing, selection may appear to be stronger. We therefore use a hierarchical model in which *s* is drawn for each country from a normal distribution, whose mean and variance we estimate in order to infer the consistency of selection. Our results particularly focus on this estimate of the mean *s* for a variant.

The migration rate, *m*, is the proportion of the country’s population swapped out for the new variant each generation. This is surely quite small, especially considering travel restrictions. We therefore set an exponential prior on *m* with mean 0.001. When the origin of a variant is strongly associated with a particular country, migration into that country is a less relevant process. We therefore removed migration (fixed *m* = 0) into the United Kingdom for B.1.1.7, into South Africa for B.1.351, and into Japan for R.1.

Because the time unit for our data is days, the estimates of *s* and *m* from the model fit must be transformed in order to be interpreted as per-generation processes. The values we report are thus all transformed as 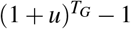, where *u* is the estimated per-day parameter and *T*_*G*_ is the generation time. The mean serial interval for SARS-CoV-2 is most likely between 4 and 7.8 days [28], so we use a normal distribution with mean 5.9 and standard deviation 1.15 for the mean generation time.

For numerical stability, we transform all frequencies to the logit scale (see Appendix A). Models were fit with Stan [29], using 4 parallel chains of length 3000, with a warm-up phase of a 1000 iterations.

### Stochastic epidemiological model

To take more detailed population dynamics into account, we use a stochastic compartmental Kermack-McKendrick-type model. In addition to susceptible (S), exposed (E), infectious (I), and removed (R) individuals, we keep track of individuals with severe disease (H), and stratify the exposed and infected populations into individuals infected with the background (wt) or the new variant (mt). The compartment of severe infections is used to model the observed delay between infection and death. The two strata are used to keep track of the new variant’s frequency in the population, and model its selective advantage (*s*).

The compartmental model keeps track of the number of individuals *S, E, I, H, R* in the disease states S, E, I, H, R, respectively. An individual starts out susceptible, and upon infection enters the exposed compartment and then becomes infectious at rate *α*. An infectious individual can either become severely infected at rate *ν*, or recover at rate *γ*. Severely infected individuals either recover or die at rate *ω*. The total population size is denoted by *N*, and we write *X* = (*S, E*_wt_, *E*_mt_, *I*_wt_, *I*_mt_, *H, R*). The transition rates *η*_*j*_(*X, t*) between the compartments are indicated by the following diagram, and the parameters are listed in Table 1.

**Table 1:** Parameters for the epidemic model for the United Kingdom (UK), the Netherlands (NL), and Japan (JP) and the D614G, B.1.1.7, B.1.351, and R.1 variants. Notes: ^a^Estimated. ^b^For the Dutch B.1.1.7 model, the initial frequency is fixed to 0. Instead, the variant is introduced due to travel from the UK. ^c^More recent estimates for sero-prevalence in the Netherlands are taken from https://www.rivm.nl/pienter-corona-studie/resultaten. ^d^The generation interval in the SEIHR model with exponentially distributed transition times is equal to 1*/α* + (*γ* + *v*)^−1^ ≈ 1*/α* + 1*/γ*. Hence, with an average incubation period of 3 days, we need an average infectious period of 4 days to get an average generation time of 7 days. ^e^By taking the probability of developing severe infection equal to 0.02 and the probability of dying from severe infection equal to 0.3, we arrive at a case fatality rate of 0.6%. Our choice is also comparable to mortality rates for ICU patients [39]. ^f^The average time between symptom onset and death is 16.5 days. After subtracting the duration of the infections period (*γ* + *v*)^−1^ ≈ 1/*γ*, we get an average duration of severe infection of 12.5 days.

| symbol | unit | value |  |  |  |  |  | description | source |
| --- | --- | --- | --- | --- | --- | --- | --- | --- | --- |
|  |  | D614G<br>UK | NL | B.1.1.7<br>UK | NL | B.1.351<br>NL | R.1<br>JP | *variant*<br>*region* |  |
| $s$ | - | 0.28 | 0.27 | 0.54 | 0.47 | 0.07 | 0.33 | selective advantage novel variant | est.<sup>a</sup> |
| $\beta_0$ | d<sup>-1</sup> | 0.81 | 0.80 | 0.39 | 0.39 | 0.33 | 0.32 | infection rate ($t < t_1$) | est. |
| $\beta_1$ | d<sup>-1</sup> | 0.15 | 0.12 | 0.24 | 0.23 | 0.25 | 0.21 | infection rate ($t_1 < t < t_2$) | est. |
| $\beta_2$ | d<sup>-1</sup> | 0.21 | 0.33 | 0.33 | 0.39 | 0.26 | - | infection rate ($t_2 < t < t_3$) | est. |
| $\beta_3$ | d<sup>-1</sup> | - | - | 0.13 | 0.22 | - | - | infection rate ($t_3 < t$) | est. |
| $t_0$ | d | 54 | 54 | 243 | 243 | 344 | 327 | initial time | - |
| $t_1$ | d | 86.6 | 82.5 | 302 | 296 | 359 | 387 | first break point $\beta$ | est. |
| $t_2$ | d | 188 | 181 | 335 | 333 | 396 | - | second break point $\beta$ | est. |
| $t_3$ | d | - | - | 370 | 354 | - | - | third break point $\beta$ | est. |
| $p_0$ | - | 0.28 | 0.37 | $3.3 \cdot 10^{-4}$ | - | 0.01 | $4.7 \cdot 10^{-3}$ | initial mutant frequency | est.<sup>b</sup> |
| $\lambda_0$ | d<sup>-1</sup> | - | - | - | $1.9 \cdot 10^{-3}$ | - | - | *per capita* infection rate due to travel | est.<sup>b</sup> |
| $r_D$ | - | 105 | 115 | 95.6 | 93.2 | 94.2 | 98.5 | overdispersion parameter death incidence | est. |
| $r_F$ | - | 72.8 | 68.3 | 81.9 | 72.1 | 75.1 | 66 | overdispersion parameter sequence data | est. |
| $\tau$ | d<sup>-1</sup> | 0.011 | 0.018 | $3.6 \cdot 10^{-3}$ | 0.024 | $3.4 \cdot 10^{-3}$ | 0.019 | overdispersion of the process noise | est. |
| $\zeta$ | - | $8.2 \cdot 10^{-6}$ | $1.7 \cdot 10^{-5}$ | $4.1 \cdot 10^{-4}$ | $3.7 \cdot 10^{-4}$ | $5.1 \cdot 10^{-3}$ | $3.8 \cdot 10^{-4}$ | initial fraction infected | est. |
| $\xi$ | - | - | - | 0.06 | 0.05 | 0.1 | 0.1 | fraction of the population immune at time $t_0$ | [31, 32]<sup>c</sup> |
| $N$ | - | 66.5 | 17.4 | 66.5 | 17.4 | 17.4 | 126 | population size (million) | - |
| $1/\alpha$ | d | 3 | . | . | . | . | . | mean duration of incubation period | [33, 34] |
| $\gamma$ | d<sup>-1</sup> | 1/4 | . | . | . | . | . | recovery rate from infectious stage | [35, 36]<sup>d</sup> |
| $\nu$ | d<sup>-1</sup> | $\gamma/50$ | . | . | . | . | . | rate of developing severe infection | [37, 38]<sup>e</sup> |
| $1/\omega$ | d | 12.5 | . | . | . | . | . | duration of severe infection | [33, 34]<sup>f</sup> |
| $\delta$ | - | 0.3 | . | . | . | . | . | probability of dying from severe infection | [37, 38, 39]<sup>e</sup> |

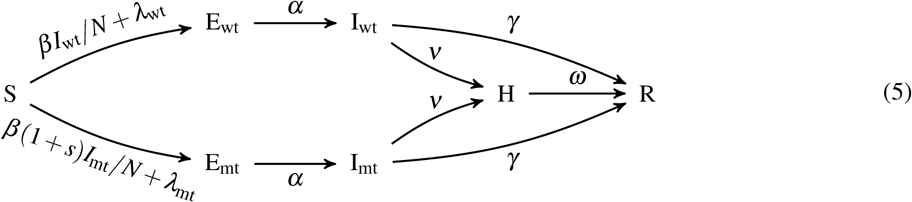

Here the indicated rates are *per capita* and should be multiplied by the size of the source compartment (e.g., *η*_H→R_(*X, t*) = *Hω*). The selective advantage of the new variant is equal to *s*; when *s* > 0, the mutant has a higher infection rate *β* (1 + *s*) than to the wild type (*β*). The other life-history traits of the virus are assumed to be identical between wild type and mutant. To model the effects of non-pharmaceutical interventions (NPI) such as lock-downs, the infection rate *β* is a smoothed, piece-wise constant function of time [30]. To account for migration, we added time-dependent terms *λ*_wt_ and *λ*_mt_ to the *per-capita* infection rate, representing the exposure of individuals in the population to SARS-CoV-2 from other regions. The precise definitions of the time-dependent parameters *β, λ*_wt_, and *λ*_mt_ are given in Appendix B.

#### Observation model

The model is fit to two data streams. The first data stream consists of weekly incidence of COVID-19 deaths *D*. For this reason, we keep track of an auxiliary accumulator variable Θ^HR^, which counts all transitions from H to R within a week. After each time the incidence is observed, the accumulator variable Θ^HR^ is set to 0. Let *δ* denote the probability that a severe infection leads to death and not recovery. To account for variability in *δ* between demographic groups or reporting errors, we use an over-dispersed negative binomial instead of a binomial or Poisson likelihood function for the observed death counts. At the time of the *n*-th observation *t*_*n*_, we then get

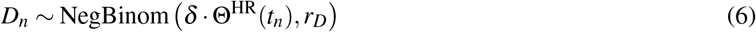

where the parameterization of the NegBinom(*ℓ, r*) distribution is such that it has mean and variance *ℓ* +*ℓ*^2^/*r*.

The second data stream consists of the number viral samples *F* that were sequenced each week, and the number sequences *F*^mt^ identified as the new variant. We assume that these sequences are collected from individuals that transition from the exposed to the infectious compartment, and hence we again define accumulator variables 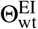 and 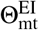 to keep track of such transitions (for wild-type and mutant infections, respectively) during the week between subsequent observation times. We define 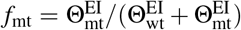 for the fraction of individuals that were infected with the new variant. To allow for over-dispersion of sampling, we use a beta-binomial likelihood function:

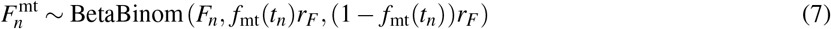

where the parameter *r*_*F*_ determines the level of over-dispersion of the sampling process.

We fit the model to the two data streams using sequential Monte-Carlo (SMC), where parameters are estimated with iterated particle filtering as described in [40]. The details of the procedure are given in Appendix B.

#### Diffusion approximation of the epidemic model

Exact simulation of the Markov jump process (MJP) that defines our stochastic epidemic model is very computationally intensive. We therefore switch to a diffusion approximation of the MJP when population sizes become large in order to do inference more efficiently. This formalism allows us to incorporate two sources of noise. The first being the process noise inherent to the MJP, which becomes negligible when the sizes of the compartments are large. We therefore introduce a second noise term that captures other origins of stochasticity that the MJP can not account for and acts on predominantly large population sizes.

As above, we denote the state of the *n*-dimensional model (where *n* = 7) by *X*^*i*^(*t*) with *i* = 1, …, *n*. The discrete, stochastic model is defined by *k* = 9 state transitions

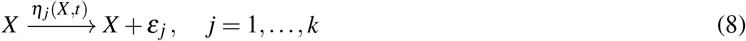

where *ε* _*j*_ ∈ ℤ^*n*^ is the increment of the *j*-th transition. For instance, the transition H → R corresponds to the increment (0,…, 0, −1, 1). Using the Kramers-Moyal expansion of the master equation, the MJP is mapped to a system of stochastic differential equations (SDE) that can be derived from the transitions *η*_*j*_ and increments *ε* _*j*_ as follows [41]

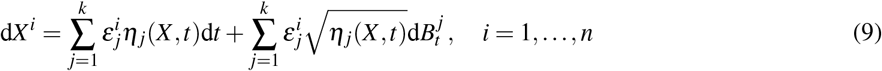

where *B*_*t*_ is a 9-dimensional Brownian motion, corresponding to the 9 transitions of the MJP in Eq. (5). The SDE in Eq. (9) is of the form d*X* = *μ*(*X, t*)d*t* + *σ*(*X, t*)d*B*_*t*_, where *μ* and *σ* describe the drift and volatility, respectively. The volatility matrix *σ*(*X, t*) encodes the intrinsic noise of the MJP, which is negligible compared to *X* when *X* is large. We therefore add a small second noise term to the system of SDEs that is proportional to *X*. After this adjustment, the SDE becomes

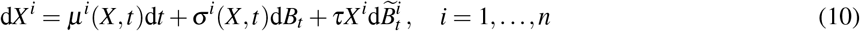

where 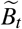 is a *n*-dimensional Brownian motion, independent of *B*_*t*_. The parameter *τ* ≪ 1 determines the magnitude of the additional noise term.

In Appendix B, we further describe in detail the algorithm used to switch from a discrete (MJP) to a continuous (SDE) model, and the way the initial condition of the system is determined.

#### Computation of confidence intervals

To compute confidence intervals for the parameter *s*, we use the profile-likelihood method. In this method, we fit the epidemic model to the data using iterated filtering, while keeping the parameter *s* fixed and record the log-likelihood. This is repeated for a sequence of values of *s*. We then fit a cubic smoothing spline through the recorded log-likelihood values, which we can maximize to obtain the maximum likelihood estimate of *s*. In addition, Wilks’ theorem allows us to compute a 95% confidence interval, by finding the values of *s* for which the log-likelihood is 1.92 units smaller than the maximum log-likelihood.

## Results

### Selective advantage of each variant in focal countries

Estimates of the selection coefficient, *s*, for each variant in a few specific countries are shown in Fig. 1. Further details from fits of the stochastic epidemiological model are shown in Figs. 2–4 and from the hierarchical population genetics model in Fig. 5. These estimates are each based on a relatively long time-series of data (generally several months, indicated in Figs. 2–4).

**Figure 1:**
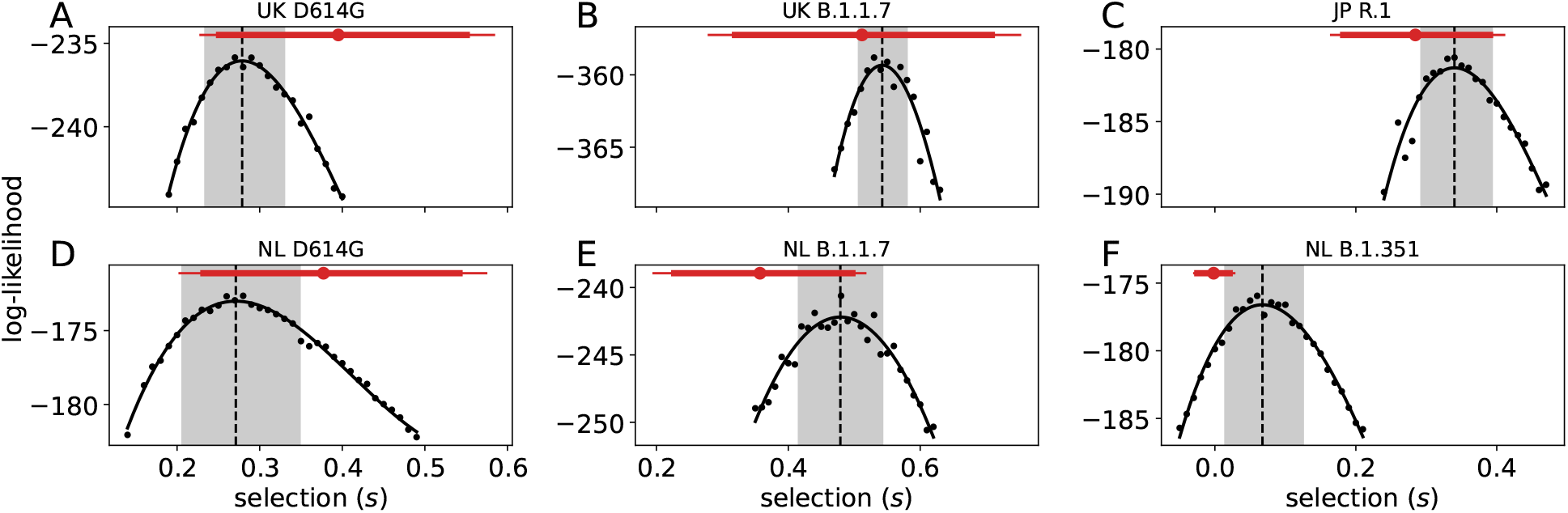
Profile likelihoods and credible intervals for the selection parameter *s* of the stochastic epidemic model for United Kingdom (UK), Netherlands (NL), and Japan (JP) and variants D614G, B.1.1.7, B.1.351, and R.1. The black dots indicate log-likelihood estimates of the fitted model with the corresponding fixed value of *s*, and the black curve is a smoothing spline through these log-likelihoods (see Methods). The dashed line shows the maximum-likelihood estimate, the gray box shows the 95% CI. The red intervals show the results of the population genetics model for these countries (cf. Fig. 5).

**Figure 2:**
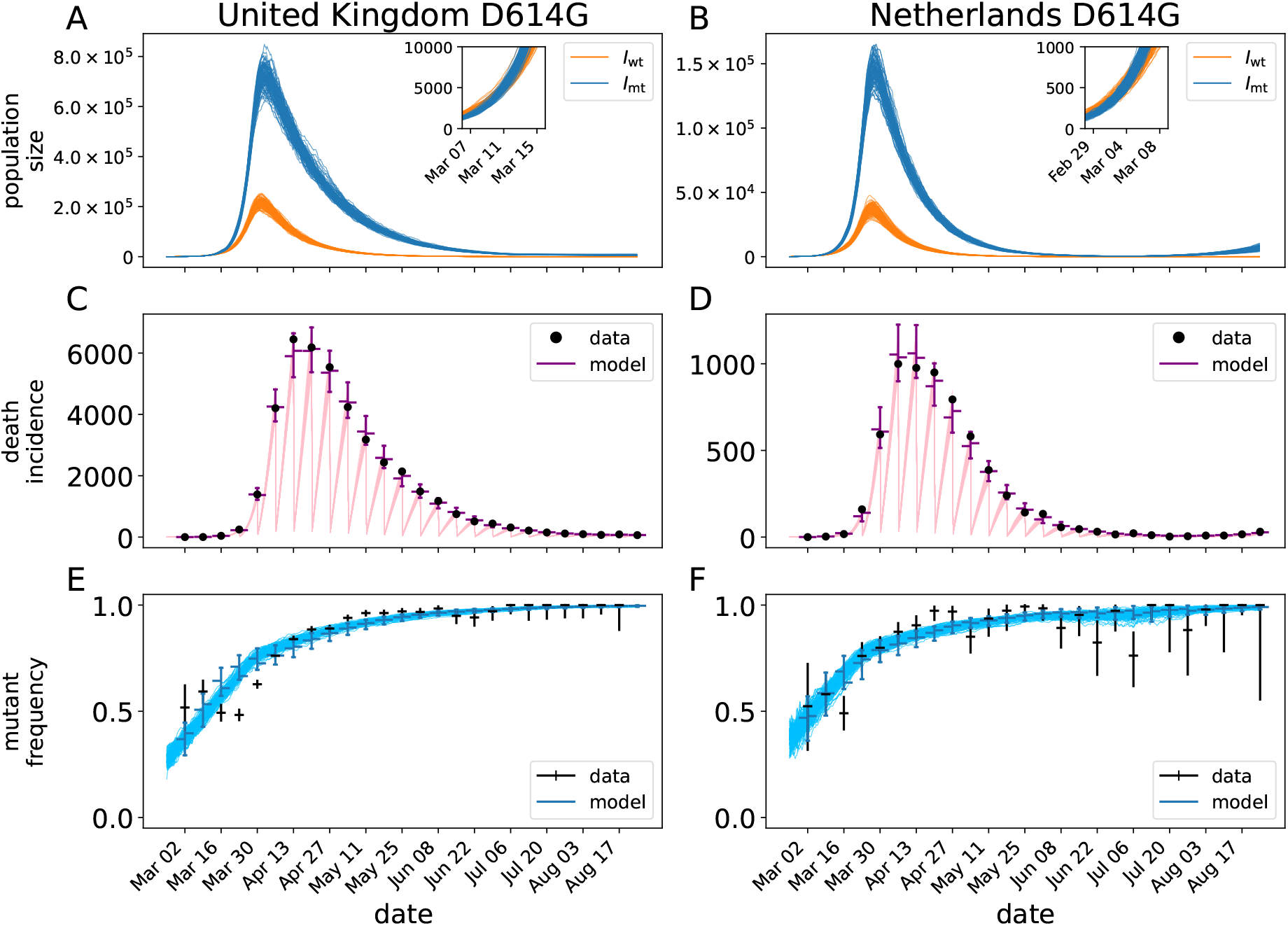
Mechanistic model fit to D614G data in the Netherlands and UK. Panels A and B show many realizations of the prevalence of the background, *I*_wt_, and D614G variant, *I*_mt_ for the maximum likelihood model fit. The insets show a close-up of the first weeks of the epidemics. Panels C and D show the number of deaths accumulated up to the week scale (black dots) and the model fits to those data. The bars around the points indicate the 95% predictive interval for the data according to the stochastic model. Panels E and F show the proportion of sequenced genomes with a glycine on position 614 of the spike protein in a given week. The blue lines are realizations of the model fit to the data. Vertical bars on the data indicate the 95% confidence intervals (CI) for the proportion based on the number of sampled genomes.

**Figure 3:**
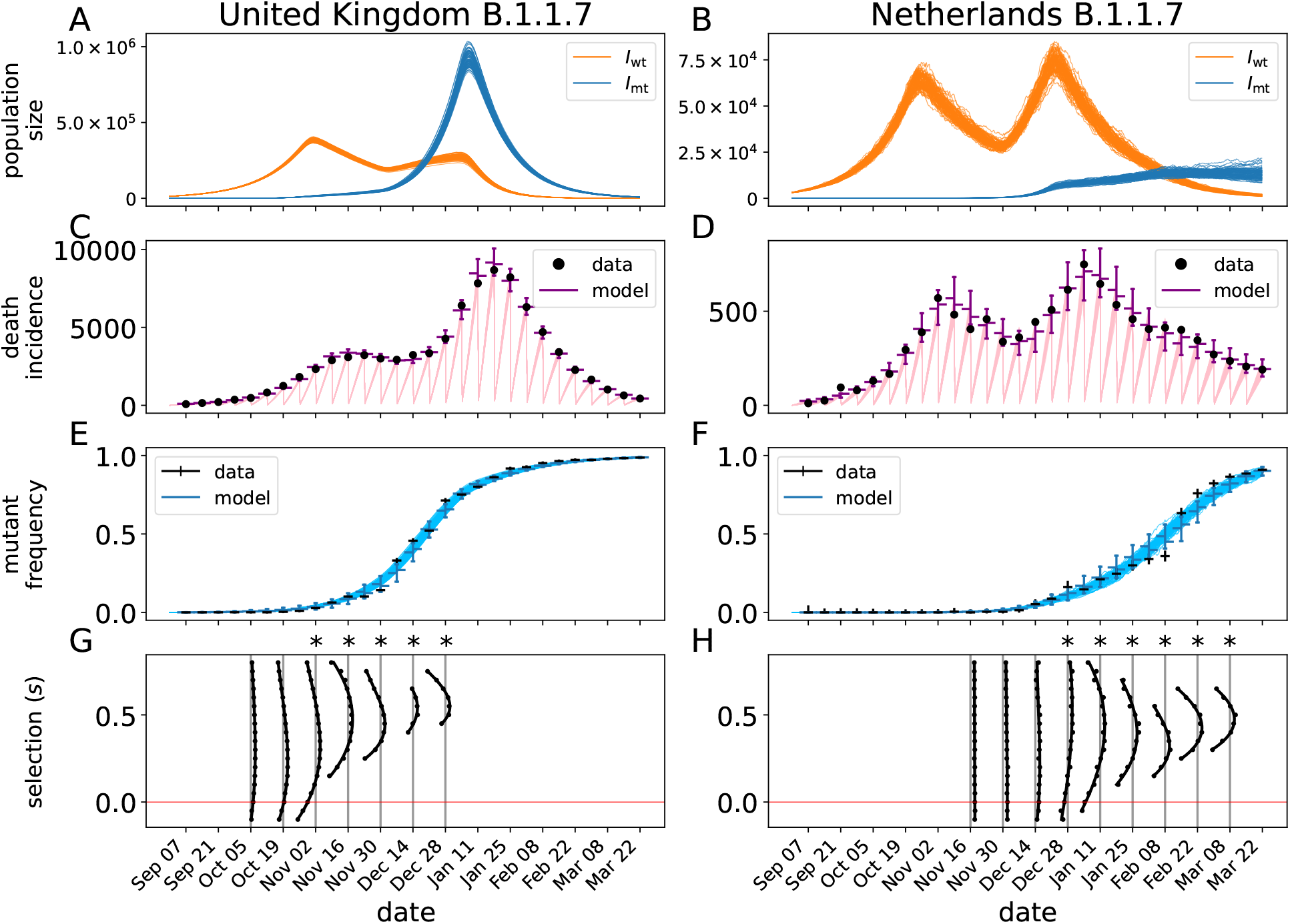
Mechanistic model fit to B.1.1.7 data in the Netherlands and UK. Panels A-H are as in Fig. 2, but now for the B.1.1.7 variant. Panels G and H show the profile-likelihood results for the time-horizon analysis at a sequence of dates. The likelihood profiles (cf. Fig. 1) are plotted vertically, where one unit in log-likelihood space corresponds to a day in calendar time. The likelihood profiles intersect with the gray vertical lines at the boundaries of the 95% CI. The horizontal red line indicates *s* = 0. The CIs marked with a star (∗) have a lower bound above *s* = 0.

**Figure 4:**
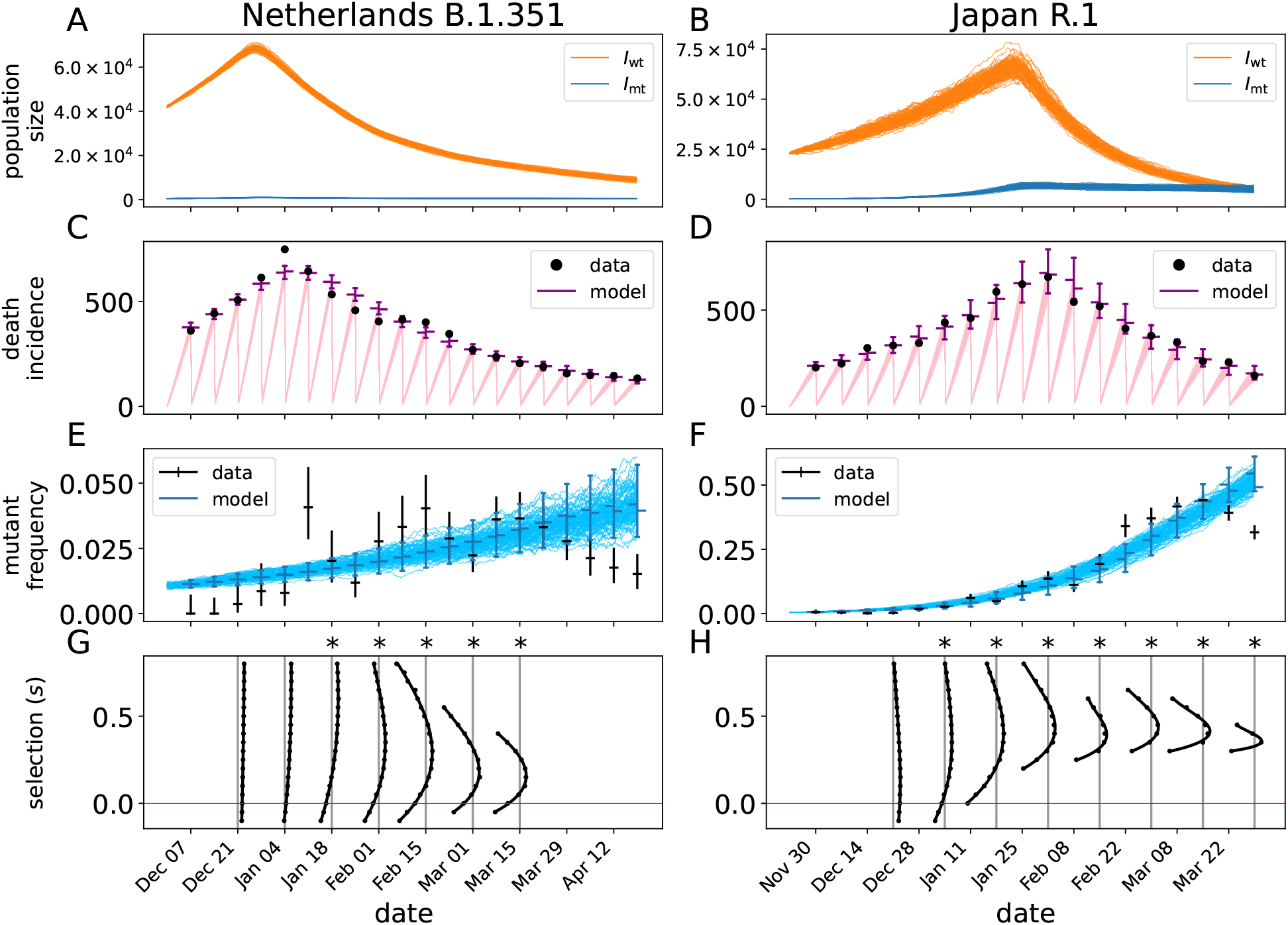
Mechanistic model fit to B.1.351 data in the Netherlands and R.1 in Japan. Panels A-F are as in Fig. 2 and panels G and H as in Fig. 3. Notice that the y-axes in panels E and F do not range from 0 to 1.

**Figure 5:**
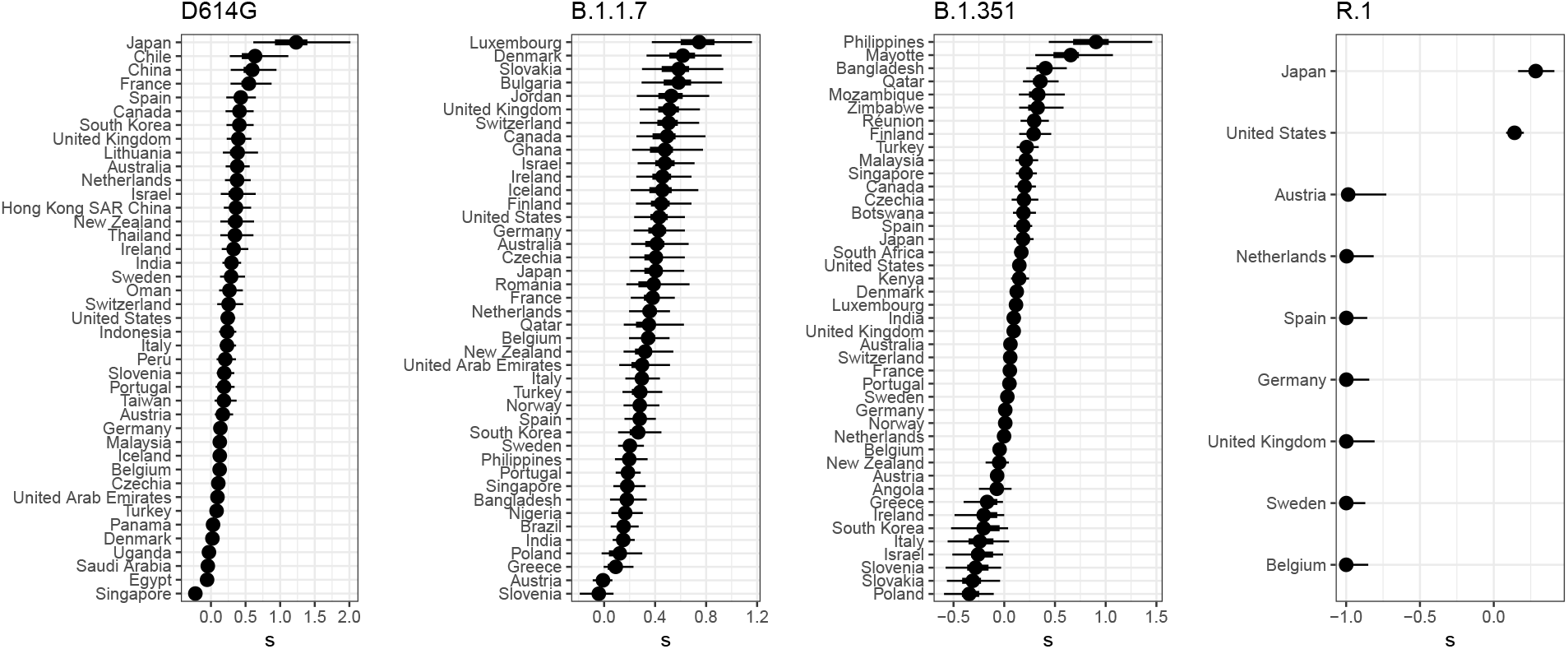
Selection coefficients for each country from the population genetic model. Results are for the final time-points shown in Figs. 2–4. Points mark the median, and thick and thin lines are 50% and 95% CIs, respectively. Corresponding estimates of migration are in Fig. S2.

We generally find overlap in the 95% CIs from the population genetics and stochastic epidemiological models, but that the estimates from the latter are substantially more precise (Fig. 1). The CIs are narrower for the stochastic epidemiological model for three main reasons. First, this model takes advantage of much more data because it incorporates case counts and deaths over time, in addition to the subset of case counts determined to be from one variant or another. Second, it fits a less-heterogeneous set of data—one country at a time. In contrast, the hierarchical population genetics model fits data from all countries simultaneously because it estimates one distribution from which is drawn a value of *s* for each country. And third, only the population genetics model incorporates uncertainty in the average generation time or serial interval. Previous work estimates an advantage of 0.1–0.3 for D614G in the UK (range across models, much broader for CIs of each model; [3]) and 0.4–0.9 for B.1.1.7 in England [2, 16]. Estimates from our stochastic epidemiological model agree and are substantially more precise (Fig. 1AB). Estimates from our population genetics model for this country do not disagree but have much larger uncertainty; across all countries the estimate is close for D614G and lower for B.1.1.7 (Fig. 7). Our search of the literature did not find previous quantitative estimates of selective advantage for B.1.351 or R.1.

The point estimates of *s* from the two methods also differ somewhat. Neither method systematically produces higher values, and to some extent there are situation-specific explanations for the differences. For example, the population genetic model estimate is probably lower for B.1.351 in the Netherlands because substantial immigration is inferred (Fig. S2), whereas migration was excluded from this fit of the stochastic epidemiological model. For R.1 in Japan, the population genetic estimate is lower because of the evidence against *s* > 0 from several other countries (Fig. 5).

Finally, the estimates of *s* from the two models are also different because they in fact represent slightly different quantities. In particular, *s* in the population genetics model is determined only by the relative growth rates of the focal and background variants (Eq. (1c)), while *s* in the stochastic epidemiological model is determined by ratio of the basic reproduction numbers. Thus, interventions that are not targeted to specific variants but affect the overall number of cases—such as wide-reaching business closures or stay-at-home orders—will mostly alter the estimate of *s* from the population genetic model, as these interventions are not explicitly modeled. We develop this idea more in Appendix C and revisit its implications in the Discussion.

### Epidemic and evolutionary dynamics in focal countries

The overall stochastic model fits to D614G and B.1.1.7 for the UK and Netherlands are shown in Fig. 2 and 3 respectively. The models provide very good fits to the data in both cases matching both the death time series and the change in proportions of the mutant variant in both countries. D614G shows a very similar pattern in the UK and Netherlands where the mutant was spreading in a way that is nearly indistinguishable from the wild-type strains for a period of several weeks in the early epidemic period. However, in both countries the model predicts that the mutant strain very quickly outpaces the wild-type strains and continues to become more relatively prevalent even when the overall prevalence is declining by orders of magnitude.

The dynamics of B.1.1.7 in the UK and Netherlands are substantially different from both D614G and each other. In both cases the mutant strain is much slower to rise, occurring over a period of months rather than weeks as was the case with D614G. In both countries, the mutant strain rises exponentially despite the large changes in the overall prevalence of COVID-19 due to changing policies and behaviors during this time. Specifically in the UK, the rise in death incidence after December 14 2020 is preceded by the rapid increase of the B.1.1.7 strain, both in frequency and absolute numbers (Fig. 3). This suggests that the interventions ongoing in the UK were sufficient to bring the background strains below threshold but not B.1.1.7.

To further substantiate this, we calculated the instantaneous effective reproduction number (*R*_*e*_) using the inferred trajectories of the stochastic model (Fig. S11). The effective reproduction number of the wild-type fluctuates abound the threshold value 1 between November and December, following the increased NPI (non-pharmaceutical intervention) initiated end October [42]. As the B.1.1.7 variant has a ∼ 50% higher reproduction number, these NPI were not sufficient for controlling the growth of the variant, leading to a doubling of the death incidence in January 2021 and the necessity of further stringent restrictions. This suggests that new variants with an increased fitness are particularly dangerous when in-place NPI are only resulting in a marginal control of the epidemic.

As the B.1.1.7 variant was most likely introduced to the Netherlands from the UK, we incorporated external forces of infection in the Netherlands (*λ*_wt_ and *λ*_mt_ in Eq. (5)) to account for this fact (see Appendix B). This process allows a source of infection in the Netherlands, governed by rate *λ*_0_ (Table 1), that is proportional to the prevalence of B.1.1.7 in the UK. We forced the migration process to zero after December 21, 2020 to account for travel restrictions from the UK to the Netherlands. Based on a sample of 100 reconstructed trajectories of the stochastic model (Fig. 3B, green curves), we estimate that at that time of the travel restriction being in place, 8468–14955 individuals were infected with the B.1.1.7 variant in the Netherlands. That is, the model suggests that the establishment of B.1.1.7 in the Netherlands was facilitated by migration from the UK, however, the major factor in the spread of B.1.1.7 in the Netherlands is its selective advantage *s*.

Not all SARS-CoV-2 variants of interest replace the background as clearly as in the case of D614G, Alpha, and Delta. To investigate how the stochastic epidemic model performs in the case of variant with a more ambiguous selective advantage, we fit the model to B.1.351 in the Netherlands, and R.1 in Japan (Fig. 4). Although the model is able to fit the death incidence data in both cases (Fig. 4C, D), the predicted mutant frequency deviates from the observed data (Fig. 4E, F). In both cases, the observed mutant frequency starts to decay after mid-March 2021, while the model prediction continues to grow. Hence the model is not capable of reproducing the observed non-monotonic mutant frequency, which is likely due, in part, to a changing background—a feature not included in the model.

### Selective advantage of each variant across all countries

For most variants in most countries, the isotonic regression method rejects the null hypothesis that the daily proportion of SARS-CoV-2 cases that are the focal variant does not increase over time (Fig. 6). The method’s computed p-values are not directly translatable into estimates of the strength of selection, but they largely correspond with estimates of *s* > 0 from the population genetics model (Figs. 5 and 6). The isotonic regression method does not distinguish whether migration can explain the change in variant frequency, rather than selection, but immigration of the new variant can not logically be the alternative explanation for a rise in frequency in all countries simultaneously.

**Figure 6:**
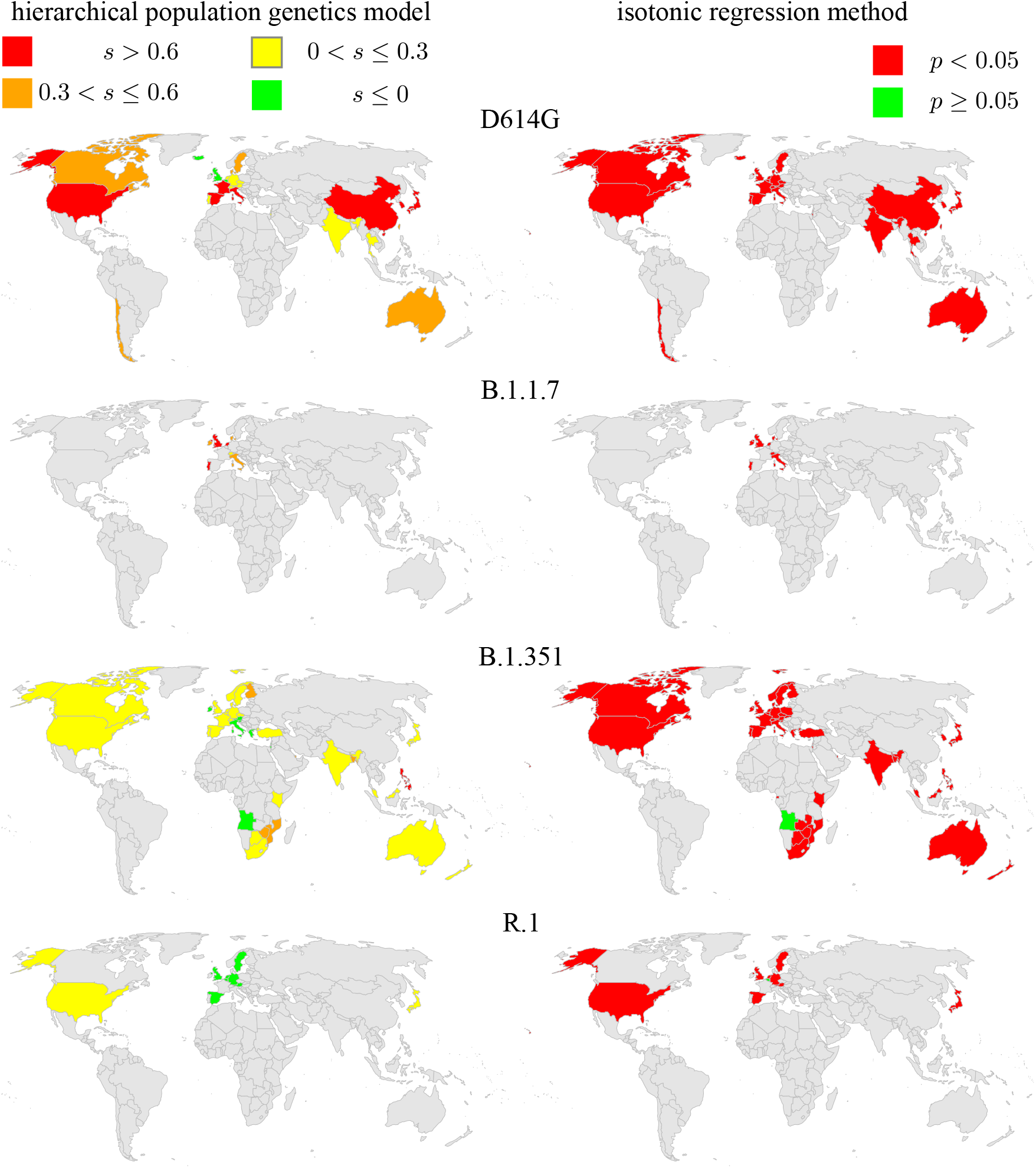
Comparison of the population genetics model and the isotonic regression method. Each variant is analyzed at the final time horizon shown in Fig. 8, and results are shown for each country with sufficient data by that time.

The hierarchical population genetics model provides an overall estimate of the mean selective advantage of each variant (Fig. 7). For D614G and B.1.1.7, these estimates are quite similar. This does not mean, however, that they are equally transmissible. The strength of selection for a variant is measured relative to all the other genotypes present over that time frame. Because B.1.1.7 emerged after D614G became globally common, the absolute fitness of B.1.1.7 is likely greater. The selective advantage of B.1.351 is less, though still positive. After its initial rise in frequency, it was overtaken by B.1.617.2 (Delta). In contrast, R.1 shows an overall strong selective disadvantage with extremely large variance among countries. It increased in frequency strongly only in Japan (Fig. S10), so the hierarchical model suggests that its rise there was due to factors other than an inherent advantage from the viral genetics, such as possibly first appearing by chance in a town or subpopulation that was experiencing an intense outbreak for other reasons.

**Figure 7:**
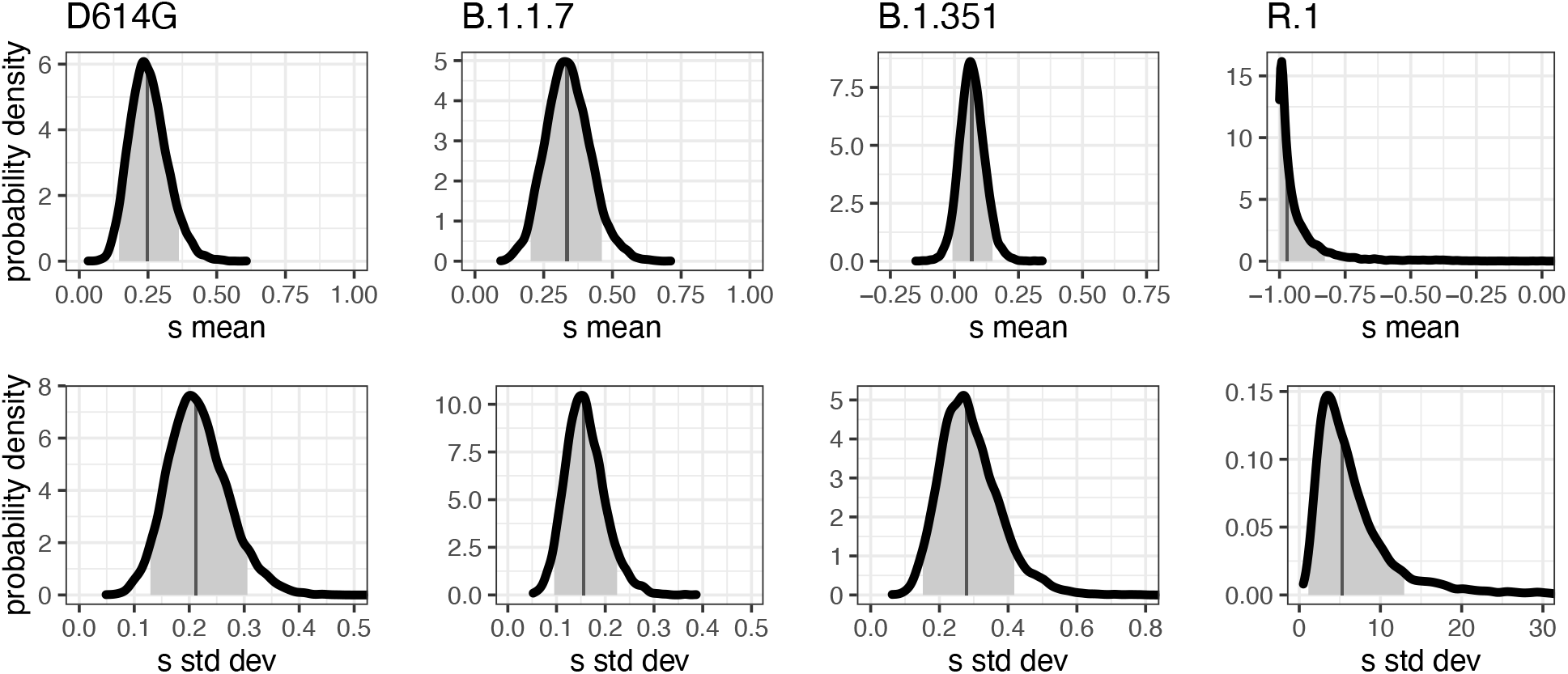
Estimated global distribution of selection coefficients for each of the four variants from the population genetics model. Our hierarchical model estimates the mean (top row) and standard deviation (bottom row) of a normal distribution from which the selection coefficient, *s*, of each country is drawn. In each panel, dark vertical lines mark the median, and the 90% CIs are shaded.

Estimates of the selection strength for each variant in each country from the population genetics model are shown in Fig. 5. For each variant, the estimates of *s* are highly heterogeneous among countries. Our model allows for a random component in country-to-country variation in *s*, but the differences in estimated *s* among the countries likely overstates the differences in actual transmission advantage. Each country surely experiences many processes that are not included in our simple model—such as superspreader events, nonrandom sampling, or waves of travellers arriving from places with different variant frequencies—and all of this heterogeneity is bundled by the model into differences in *s* (and *m*, which is constrained to be small). Furthermore, strong selection for one variant in one country does not necessarily correspond to strong selection for another variant in that country, suggesting that factors beyond country-level covariates underlie the overall heterogeneity.

In estimating the selective advantage of each variant, our population genetics model allows for a contribution of migration in elevating the variant frequencies. Because selection and migration to some extent provide alternative explanations for change in variant frequency, we find some negative correlation between these two processes (Figs. S3–S6). The estimates of migration are not particularly distinguishable among countries (Fig. S2), but estimates of selection nevertheless show clear differences among countries (Fig. 5). We thus conclude that the selective effect of a variant can be estimated even allowing for a reasonable amount of migration.

### Time to detect a selective advantage of a variant

Ideally we would want to know that a new variant has a selective advantage as soon as possible to give as much time as possible to implement interventions. To see how rapidly our methods could reliably detect a selective advantage, we limited the data for each variant to a series of past time horizons.

Both the stochastic epidemiological and population genetics model identify *s* > 0 for B.1.1.7 in the UK by mid-November (Fig. 8, Fig. 3G). This variant was slow to spread to other countries (or at least, to be detected elsewhere), so the hierarchical model does not apply until late December, and at that time it does not much change the estimate of mean *s*. By mid-January, this variant’s advantage was also detectable in the Netherlands (Fig. 3H). However, the the estimate of *s* appeared to be declining from late January to early February likely due to a short period where the mutant frequency seemed to plateau. While the full run of the data was very consistent with the model, this aberration could be easily over-interpreted in a real-time environment, highlighting the utility of both detailed, country-specific models with broader multi-region approaches. Similarly, the selective advantage of D614G was detectable once there were 2500 cases globally, and the estimate of *s* remained approximately the same over the next month as the number of cases increased to 12500 (Fig. 8).

**Figure 8:**
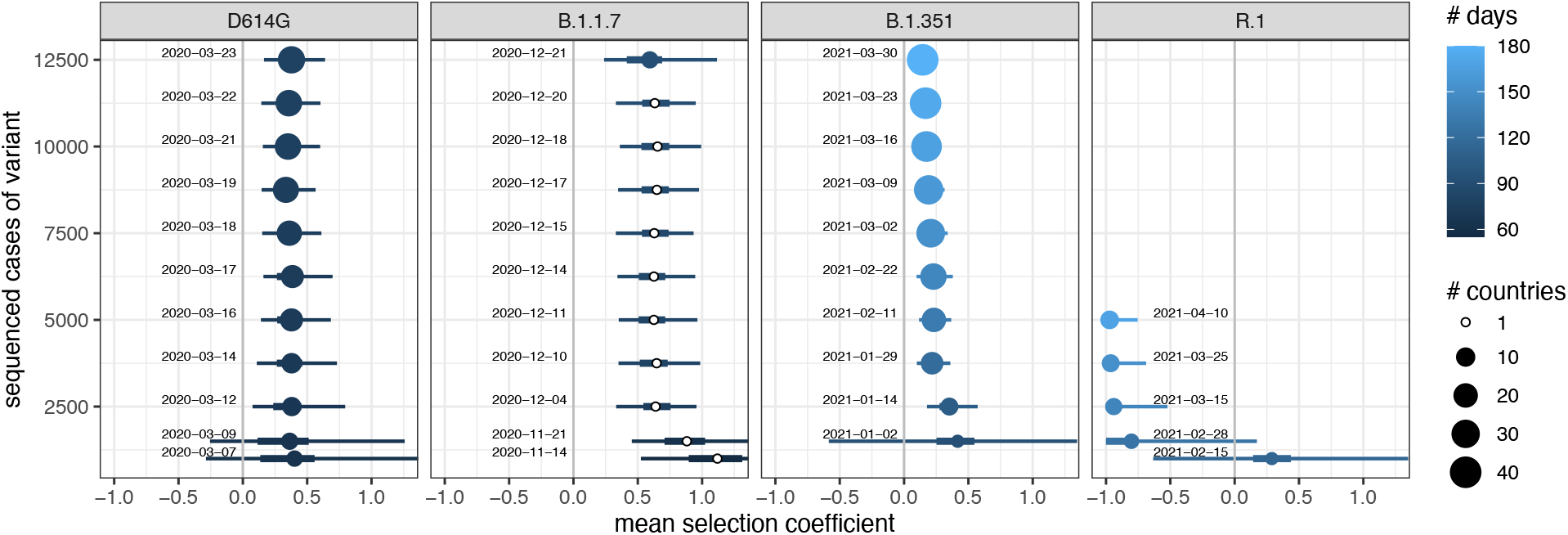
Estimates of the global selection coefficient, from the hierarchical population genetics model, for each variant for differing amounts of data. (When only one country was present in the data, the non-hierarchical equivalent model was fit instead.) Dates show the first day on which that corresponding number of cases of that variant was first reached globally; they are the time horizon to which the data were cut back for that estimate. The number of days of data is measured from the time horizon back to a first day for each variant: 2020-01-04 for D614G, 2020-09-20 for B.1.1.7, 2020-10-01 for B.1.351, and 2020-10-24 for R.1. Points mark the median, and thick and thin lines are 50% and 95% CIs, respectively.

In contrast, B.1.351 was slower to increase in global number of cases and reached many countries during its initial expansion. During this expansion, the hierarchical model detected *s* > 0 after 2500 total cases, but the estimate of mean *s* then declined toward zero over the next two months (Fig. 8), perhaps due to the rise of B.1.617.2 (Delta) which emerged later but rapidly came to dominate cases globally. Using the stochastic epidemic model, we could confirm this pattern in the Netherlands (Fig. 4G). The lower bound of the 95% CI crosses *s* = 0 after January 4 2021, and slightly increases until February 15, but then starts to decrease as the B.1.351 frequency plateaus.

The variant R.1 shows yet a different pattern. It initially rose in frequency in Japan, and the stochastic epidemiological model found *s* confidently greater than zero by early January. The first 1000 cases globally were not reached until mid-February, at which point the strikingly different trajectories in different countries (Japan and United States versus Austria) induced enormous uncertainty in the estimate of mean *s* for the hierarchical model (Fig. 8). Over the next two months, the variant reached more countries but did not substantially increase in frequency within them, leading to a strongly negative estimate of mean *s*.

Overall, both the population genetic and stochastic models were in agreement on when a variant could be determined to be a concern. However, the population genetic model’s focus on the global distribution of selection effects was able to avoid wrongly concluding that R.1 was a variant of concern due to the anomalous rise of that variant in Japan.

## Discussion

### Methods for genomic surveillance

We have illustrated three different approaches to measuring selection effects from the global SARS-CoV-2 genetic sequence data. Our analyses point to strong selection favoring D614G and B.1.1.7, some selection favoring B.1.351, and not a global advantage for R.1. SARS-CoV-2 is rapidly adapting to its new human hosts, and variants with elevated contagiousness will surely continue to emerge as the virus continues to adapt. Integrating molecular epidemiology surveillance into SARS-CoV-2 pipelines is essential for not only monitoring the emergence of new strains, but for establishing an early warning system to monitor for escape mutations in the era of vaccine roll-out. A central question in any modelling endeavor is how much detail is required to accurately address the problem in question. In the last year, a large number of models have been developed to study various aspects of the SARS-CoV-2 pandemic, ranging from very simple [33] to quite detailed [16]. We developed both simple and more complex approaches and showed how each has its own strengths and weaknesses, and how they can fit into an expanded molecular epidemiology surveillance system.

The isotonic regression method is easy to compute and based on the very straightforward premise that a consistent selective advantage should produce a continually increasing frequency of the new variant in all countries where it has been observed. However, because the method is based on a hypothesis-testing framework, there is no way to quantify the strength of selection relative to the background strains. Given the rapid pace of COVID-19 variant epidemiology research and the interest of general audiences in the results of that research, scientists need to be realistic about the possibility of p-values being wrongly interpreted as a measure of selection strength. Additionally, this method identified a significant result for most variants in most countries we analyzed, likely because we had already chosen variants of interest based on their initial rise in frequency. We believe that a non-parametric regression-based approach is, nevertheless, very useful for rapidly evaluating evidence of selection potentially in large-scale molecular surveillance pipelines.

Our population genetics model is more mechanistically explicit than the isotonic regression approach and consequently yields an estimate of the selection effect. The model also allowed us to integrate a simple migration process and to jointly estimate the parameters of selection and migration. The population genetic model is also simple enough that it was coded in a popular statistical language and fit to the global data in a matter of hours on a standard laptop computer. Its framework to estimate country-level selection effects shaped by an overall global distribution makes it potentially quite useful for general molecular surveillance purposes. For example, the rise of R.1 in Japan was attributed to a selection advantage by our stochastic epidemiological model likely because it did not include extremely detailed local effects (e.g., perhaps R.1 arose in a portion of a contact network with high tranmission for other reasons), whereas the hierarchical population genetics model weighed evidence from other countries to conclude that R.1 was not a variant likely to spread widely. One limitation is that this model includes no epidemiological structure, making it impossible to distinguish fitness advantages due to different mechanisms, e.g., greater contagiousness versus longer infectious period. The primary weakness of this model is that it is deterministic, lacking drift. Not accounting for random fluctuations in the underlying populations is potentially a problem when the goal is estimating selection effects in near real-time in order to give warning before a new variant becomes widespread.

Our stochastic epidemiological model explicitly includes effects that could produce changes in variant frequency by chance alone, in addition to selection and migration. At one level, we include noise expected in a typical model of homogeneous mixing at the country level. However in reality, transmission is occurring at much smaller local scales that can lead to sudden jumps in both the number of cases and number of observed variants, so we allow an additional noise term that makes the method less sensitive to misspecifications such as an over-simplified population structure. Such a noise term could potentially also be included in methods that allow for more efficient Bayesian inference with stochastic models [43]. Despite being more complex and using additional data, we found that the stochastic epidemiological model was in agreement with the population genetic model, suggesting that the population genetic model is a reasonable balance between computability and accuracy.

Several studies using other methods have similarly found D614G and B.1.1.7 to each have a selective advantage over other variants circulating contemporaneously [14, 27, 3, 16, 2]. This includes phylodynamic methods, which explicitly consider the evolutionary relationships among all samples—not merely the variant name for each sample, as we use here—and fit an epidemiological model to these phylogenetic data. Such methods draw more power from the data, but at such a computational expense that only small datasets can be analyzed. A different type of phylogenetic analysis found no support for a selective advantage of any of the variants they tested, including D614G [44], presumably because their statistical test required the repeated emergence of a variant in order to draw any power. Although phylogenetic replication is an appropriate requirement in many situations, it is too conservative for identifying variants of concern on the timescale at which they emerge. Instead, to test for a selective advantage of variants that have arisen only once, power can be obtained from fitting explicitly epidemiological models within one location (e.g., our stochastic model, and others [27, 3, 16, 2]) or looking for consistent effects in multiple locations with largely-distinct conditions (e.g., our isotonic regression and population genetic models, and Korber et al. [14]).

### Estimating and interpreting selective advantage

One important limitation of all our approaches is that they look for an advantage of a given variant over whatever other variants are circulating at the time, and the background of other variants is constantly changing. This means, for example, that the fitness advantage estimated for B.1.1.7 is relative to a background that consists mostly of D614G. That is, the fitness of B.1.1.7 exceeds that of D614G, which itself exceeds the original genotype. Similarly, B.1.351 initially exhibited an advantage, but this was later reduced probably due the rise of B.1.617.2. This complication of a shifting fitness landscape is reduced in our analyses that model effects only over early time horizons for each variant, but it is not eliminated. A possible solution would be to develop models that track multiple, simultaneously-competing variants, with the fitness of each variant measured against a fixed reference strain [45]. An additional complication is that the fitness landscape is changed not only by the presence of other variants, but also potentially by changes in host immunology and vaccination. And of course each variant is not a fixed entity: they are given discrete names for convenience, but each viral lineage continues to accumulate new mutations.

Biased sampling will always be a potential problem when taking advantage of haphazardly gathered data. All of our models (and most other published models) make the assumption that genomes are selected at random from the set of all possible cases. If, for example, samples were sequenced specifically because they were in contact with someone that was known to be infected by the variant under study, the data may be biased toward over-estimating the spread and hence selective advantage of the new variant. There is almost certainly some bias from the non-random processes by which samples are obtained and sequenced; however, we believe that our results are still overall valid for three reasons. First, it is unlikely that the same level of bias from non-random sampling would occur in each country to produce a similar pattern in each country; that is, countries represent semi-independent systems. Second, the evidence for selection effects includes parts of the time series before people were concerned about the spread of new variants, and, therefore, were unlikely to preferentially sequence the new variants. Third, the UK has put effort into developing a representative sample of SARS-CoV-2 genomes in their country and the estimates for the selection effects in the UK for D614G and B.1.1.7 are very close and slightly above the population average for these lineages, while being only slightly below average for B.1.351.

Increasing vaccination rates will have several effects on estimates of variant fitness. A new form of sampling bias is possible, if breakthrough cases are preferentially sequenced. If vaccination leads to fewer cases overall, early estimates particularly from the regression and population genetic models will have greater uncertainty. Eventually we might expect to see variants that specifically evade a vaccine—these could perhaps be identified early by finding a pattern in which a variant has a higher selective advantage in places with higher vaccination rates, suggesting a mixed-effects modeling approach instead of the current random-effects approach.

In both our population genetic and stochastic epidemiological models, we use *s* to denote the selective advantage of the focal variant, and the definition of *s* in each model makes intuitive sense as a transmission advantage. However, our model analysis shows that a news story reporting that a new variant is, say, ‘30% more transmissible’ would translate to different meanings in the two models depending on the value of *R*_*e*_ (Fig. S1 and Appendix C). One important conclusion is that an intervention lowering the total proportion of people who are infected causes the transmission advantage to be underestimated by the population genetics model because this advantage scales with the effective reproduction number (see Appendix C). Although this type of difference may be difficult to communicate to a general audience, it must be accounted for in scientific studies that quantify and compare fitnesses of different variants, particularly when these are estimated by different methods. However, such comparisons are additionally fraught because continual changes in the environment make it likely impossible to define a single, true, consistent value of fitness for each variant.

The emergence of new variants with increased contagiousness or resistance mutations has significant implications for control of COVID-19, especially given that very few countries have been able to use NPIs alone to bring the viral growth rate sub-critical for extended periods of time. This situation is challenging as elevated contagiousness narrows the range under which vaccination programs can eliminate the virus, and it also opens up the possibility of escape mutations allowing infection among vaccinated persons. Integrating modeling into surveillance systems will help facilitate early-warning systems and improve our ability to design both pharmaceutical and non-pharmaceutical interventions that can stop the spread of COVID-19.

## Data Availability

All data and scripts are publicly available

https://github.com/eeg-lanl/sarscov2-selection

## Acknowledgements

Portions of this work were done under the auspices of the U.S. Department of Energy under contract 89233218CNA000001 and supported by National Institutes of Health (www.nih.gov) grants P01-AI131365, R01-OD011095, and R01-AI028433 (CHvD). RK, NH, and ERS were funded by the US National Science Foundation RAPID grant PHY-2031756. Research presented in this article was supported by the Laboratory Directed Research and Development program of Los Alamos National Laboratory under project numbers 20210528CR and 20210887ER.

## Literature Cited

[1] Tegally, H. et al. Detection of a SARS-CoV-2 variant of concern in South Africa. Nature 592, 438–443 (2021).

[2] Volz, E. et al. Transmission of SARS-CoV-2 lineage B.1.1.7 in England: Insights from linking epidemiological and genetic data. medRxiv (2021).

[3] Volz, E. et al. Evaluating the Effects of SARS-CoV-2 Spike Mutation D614G on Transmissibility and Pathogenicity. Cell 184, 64–75 (2021).

[4] Challen, R. et al. Early epidemiological signatures of novel SARS-CoV-2 variants: establishment of B.1.617.2 in England. medRxiv (2021).

[5] Dearlove, B. et al. A SARS-CoV-2 vaccine candidate would likely match all currently circulating variants. Proceedings of the National Academy of Sciences 117, 23652–23662 (2020).

[6] Singh, J., Rahman, S. A., Ehtesham, N. Z., Hira, S. & Hasnain, S. E. SARS-CoV-2 variants of concern are emerging in India. Nature Medicine 1–3 (2021).

[7] Fontanet, A. et al. SARS-CoV-2 variants and ending the COVID-19 pandemic. Lancet (2021).

[8] McCarthy, K. R. et al. Recurrent deletions in the SARS-CoV-2 spike glycoprotein drive antibody escape. Science (2021).

[9] Wibmer, C. K. et al. SARS-CoV-2 501Y.V2 escapes neutralization by South African COVID-19 donor plasma. bioRxiv (2021).

[10] Weisblum, Y. et al. Escape from neutralizing antibodies by SARS-CoV-2 spike protein variants. eLife 9, e61312 (2020).

[11] Cobey, S., Larremore, D. B., Grad, Y. H. & Lipsitch, M. Concerns about SARS-CoV-2 evolution should not hold back efforts to expand vaccination. Nat Rev Immunol 21, 330–335 (2021).

[12] Gerrish, P. J. et al. How unequal vaccine distribution promotes the evolution of vaccine escape. Available at SSRN 3827009 (2021).

[13] Grubaugh, N. D. et al. Tracking virus outbreaks in the twenty-first century. Nat Microbiol 4, 10–19 (2019).

[14] Korber, B. et al. Tracking Changes in SARS-CoV-2 Spike: Evidence that D614G Increases Infectivity of the COVID-19 Virus. Cell 182, 812–827 (2020).

[15] Yurkovetskiy, L. et al. Structural and Functional Analysis of the D614G SARS-CoV-2 Spike Protein Variant. Cell 183, 739–751 (2020).

[16] Davies, N. G. et al. Estimated transmissibility and impact of SARS-CoV-2 lineage B.1.1.7 in England. Science (2021).

[17] Davies, N. G. et al. Increased mortality in community-tested cases of SARS-CoV-2 lineage B.1.1.7. Nature (2021).

[18] Frampton, D. et al. Genomic characteristics and clinical effect of the emergent SARS-CoV-2 B.1.1.7 lineage in London, UK: a whole-genome sequencing and hospital-based cohort study. Lancet Infect Dis 21, 1246–1256 (2021).

[19] Twohig, K. A. et al. Hospital admission and emergency care attendance risk for SARS-CoV-2 Delta (b.1.617.2) compared with Alpha (b.1.1.7) variants of concern: a cohort study. The Lancet Infectious Diseases (2021).

[20] Mlcochova, P. et al. SARS-CoV-2 B.1.617.2 Delta variant replication, sensitivity to neutralising antibodies and vaccine breakthrough. bioRxiv (2021).

[21] Hirotsu, Y. & Omata, M. Detection of R.1 lineage severe acute respiratory syndrome coronavirus 2 (SARS-CoV-2) with spike protein W152L/E484K/G769V mutations in Japan. PLoS Pathogens 17, e1009619 (2021).

[22] Dong, E., Du, H. & Gardner, L. An interactive web-based dashboard to track COVID-19 in real time. The Lancet Infectious Diseases 20, 533–534 (2020).

[23] Elbe, S. & Buckland-Merrett, G. Data, disease and diplomacy: GISAID’s innovative contribution to global health. Glob Chall 1, 33–46 (2017).

[24] Global Initiative on Sharing All Influenza Data. http://www.gisaid.org/ (2008).

[25] COVID-19 Viral Genome Analysis Pipeline. https://cov.lanl.gov (2020).

[26] Liao, X. & Meyer, M. C. cgam: An r package for the constrained generalized additive model. Journal of Statistical Software, Articles 89, 1–24 (2019).

[27] Chen, C. et al. Quantification of the spread of SARS-CoV-2 variant B.1.1.7 in Switzerland. medRxiv (2021).

[28] Ali, S. T. et al. Serial interval of SARS-CoV-2 was shortened over time by nonpharmaceutical interventions. Science 369, 1106–1109 (2020).

[29] Carpenter, B. et al. Stan: A probabilistic programming language. J. Stat. Softw. 76, 1–32 (2017).

[30] Rozhnova, G. et al. Model-based evaluation of school-and non-school-related measures to control the COVID-19 pandemic. Nature Communications 12, 1614 (2021).

[31] Ward, H. et al. Antibody prevalence for SARS-CoV-2 following the peak of the pandemic in england: REACT2 study in 100,000 adults. medRxiv (2020).

[32] Vos, E. R. A. et al. Nationwide seroprevalence of SARS-CoV-2 and identification of risk factors in the general population of the Netherlands during the first epidemic wave. J Epidemiol Community Health (2020).

[33] Sanche, S. et al. High Contagiousness and Rapid Spread of Severe Acute Respiratory Syndrome Coronavirus 2. Emerg Infect Dis 26, 1470–1477 (2020).

[34] Zhou, F. et al. Clinical course and risk factors for mortality of adult inpatients with COVID-19 in Wuhan, China: a retrospective cohort study. Lancet 395, 1054–1062 (2020).

[35] Ali, S. T. et al. Serial interval of SARS-CoV-2 was shortened over time by nonpharmaceutical interventions. Science 369, 1106–1109 (2020).

[36] Lavezzo, E. et al. Suppression of a SARS-CoV-2 outbreak in the Italian municipality of Vo’. Nature 584, 425–429 (2020).

[37] Wu, J. T. et al. Estimating clinical severity of COVID-19 from the transmission dynamics in Wuhan, China. Nat Med 26, 506–510 (2020).

[38] Verity, R. et al. Estimates of the severity of coronavirus disease 2019: a model-based analysis. Lancet Infect Dis 20, 669–677 (2020).

[39] Grasselli, G. et al. Baseline Characteristics and Outcomes of 1591 Patients Infected With SARS-CoV-2 Admitted to ICUs of the Lombardy Region, Italy. JAMA 323, 1574–1581 (2020).

[40] Ionides, E. L., Nguyen, D., Atchadé, Y., Stoev, S. & King, A. A. Inference for dynamic and latent variable models via iterated, perturbed Bayes maps. Proceedings of the National Academy of Sciences 112, 719–724 (2015).

[41] van Kampen, N. G. Stochastic processes in physics and chemistry. Elsevier, Amsterdam, 3rd edition (2007).

[42] Hale, T., Webster, S., Petherick, A., Phillips, T. & Kira, B. Oxford COVID-19 Government Response Tracker (2020).

[43] Fintzi, J. et al. Using multiple data streams to estimate and forecast SARS-CoV-2 transmission dynamics, with application to the virus spread in Orange County, California (2020).

[44] van Dorp, L. et al. No evidence for increased transmissibility from recurrent mutations in SARS-CoV-2. Nature Communications 11, 5986 (2020).

[45] Annavajhala, M. K. et al. Emergence and expansion of the sars-cov-2 variant b.1.526 identified in new york. medRxiv (2021).

