## Appendices and Supplementary Figures for "Estimating the strength of selection for new SARS-CoV-2 variants"

### Supporting Information

Christiaan H. van Dorp[1,*], Emma E. Goldberg[1,2,*], Nick Hengartner[1,2], Ruian Ke[1,2], and Ethan O. Romero-Severson[1,2,†]

[1]Theoretical Biology and Biophysics (T-6), Los Alamos National Laboratory, Los Alamos NM, USA

[2]New Mexico Consortium, Los Alamos NM, USA

[*]These authors contributed equally to this work

September 24, 2021

#### Appendix A: Population genetics model

##### *Variant frequencies over time*

Here we derive the equation for the mutant frequency $p_n$ as a function of the generation $n$ under the population genetics model with both selection and migration (Eq 4). In the main text, we derived that the new variant's frequency $p'$ in the next generation is

$$p' = \frac{(1+s)p + (1-p)m}{1+sp}. \tag{A-1}$$

We can write this slightly differently as a Möbius transformation of $p$:

$$p' = \frac{(1+s-m)p+m}{sp+1} \equiv \begin{pmatrix} 1+s-m & m \\ s & 1 \end{pmatrix} \cdot p. \tag{A-2}$$

In general a Möbius transformation has the form

$$\begin{pmatrix} a & b \\ c & d \end{pmatrix} \cdot x \equiv \frac{ax+b}{cx+d}, \tag{A-3}$$

and it has the nice property that $A \cdot (B \cdot x) = (AB) \cdot x$ for two matrices $A$ and $B$ (see e.g., [1]). Hence, in order to find the variant frequency in the $n$-th generation, we need the $n$-th matrix power of

$$M = \begin{pmatrix} 1+s-m & m \\ s & 1 \end{pmatrix}. \tag{A-4}$$

For this we have to diagonalize $M$. The eigenvalues of $M$ are equal to $1+s$ and $1-m$. Then eigenvectors are given by $(1,1)^T$ and $(m,-s)^T$. Hence, we can write $M = U\Lambda U^{-1}$ where

$$U = \begin{pmatrix} 1 & m \\ 1 & -s \end{pmatrix} \quad \text{and} \quad \Lambda = \begin{pmatrix} 1+s & 0 \\ 0 & 1-m \end{pmatrix}. \tag{A-5}$$

Then $M^n \cdot p_0 = U\Lambda^n U^{-1} \cdot p_0$, which is equal to

$$p_n = \frac{[s(1+s)^n + m(1-m)^n]p_0 + m[(1+s)^n - (1-m)^n]}{s[(1+s)^n - (1-m)^n]p_0 + [m(1+s)^n + s(1-m)^n]}. \tag{A-6}$$

##### *Transformation to the logit scale*

To avoid numerical issues during inference, we transform the mutant frequency to the logit-scale, and re-parameterize the model. Let $R = \left(\begin{smallmatrix} 1 & 0 \\ -1 & 1 \end{smallmatrix}\right)$. Then

$$\text{logit}(x) = \log(R \cdot x). \tag{A-7}$$

Let $r_0 = R \cdot p_0$ and $r_n = R \cdot p_n$. Then

$$r_n = (RU\Lambda^n U^{-1} R^{-1}) \cdot r_0, \tag{A-8}$$

and we have $RU = \begin{pmatrix} 1 & m \\ 0 & -(m+s) \end{pmatrix}$. Hence,

$$\left(RU\Lambda^n (RU)^{-1}\right) = \begin{pmatrix} (1+s)^n & \frac{m}{m+s}\left[(1+s)^n - (1-m)^n\right] \\ 0 & (1-m)^n \end{pmatrix} \tag{A-9}$$

and therefore

$$r_n = \left(\frac{1+s}{1-m}\right)^n r_0 + \frac{m}{m+s}\left[\left(\frac{1+s}{1-m}\right)^n - 1\right]. \tag{A-10}$$

This equation has a removable singularity at $s = -m$. To see this, we write

$$\frac{m}{m+s}\left[\left(\frac{1+s}{1-m}\right)^n - 1\right] = \frac{m}{1-m}\frac{\left(\frac{1+s}{1-m}\right)^n - 1}{\frac{1+s}{1-m} - 1} = \frac{m}{1-m}\sum_{k=0}^{n-1}\left(\frac{1+s}{1-m}\right)^k. \tag{A-11}$$

Now we define $\sigma = \frac{1+s}{1-m}$ and $\mu = m/(1-m)$. Then we get the following re-parameterized model:

$$r_n = \sigma^n r_0 + \mu \frac{1-\sigma^n}{1-\sigma}. \tag{A-12}$$

#### Appendix B: Stochastic epidemiological model

***Time-dependent infection rate***

To model the effects of government restrictions such as lock-downs, the infection rate $\beta$ is a (smoothed) piece-wise constant function of time. We allow for $n \in \{3,4\}$ epidemic stages to model e.g. unrestricted spread, lockdown, and relaxation of the lockdown [2]. We let $\beta$ vary smoothly between the epidemic stages, thereby allowing for an uptake period for (release of) restrictions. More precisely, $\beta = \beta(t)$ is defined as

$$\begin{aligned} \beta(t) = {} & \beta_0 (1 - H_\upsilon(t - t_1)) \\ & + \sum_{i=1}^{n-2} \beta_i \cdot H_\upsilon(t - t_i)(1 - H_\upsilon(t - t_{i+1})) \\ & + \beta_{n-1} \cdot H_\upsilon(t - t_{n-1}) \end{aligned} \tag{B-1}$$

where $H_\upsilon$ is a smoothed Heaviside function defined by $H_\upsilon(t) = (1 + \exp(-t/\upsilon))^{-1}$, with parameter $\upsilon$ determining the duration of the transition period between different epidemic stages. The time-varying infection rate $\beta$ is illustrated in Fig. S11A. To determine the parameter $\upsilon$, we require that $H_\upsilon(\Delta t/2) = 95\%$, where $\Delta t$ is the length of the uptake period. Solving for $\upsilon$, we find $\upsilon = \Delta t/(2 \cdot \mathrm{logit}(0.95))$. We set $\Delta t = 7$ days, hence the uptake period takes a week.

In the case of variant B.1.1.7 in the Netherlands, we added an external force of infection, $\lambda_{\mathrm{wt}}$ and $\lambda_{\mathrm{mt}}$ for wild-type and mutant respectively, to account for contacts with infectious individuals from the UK. Because the number of infected individuals the UK varied over time, the external infection forces of infection $\lambda_{\mathrm{wt}}$ and $\lambda_{\mathrm{mt}}$ depend on time as follows

$$\lambda_{\mathrm{wt}}(t) = \lambda_0 \mathbb{E}[I_{\mathrm{wt}}(t)/N(t)]\,, \quad \lambda_{\mathrm{mt}}(t) = \lambda_0 \mathbb{E}[I_{\mathrm{mt}}(t)/N(t)]\,. \tag{B-2}$$

Here, the trajectories $I_{\mathrm{wt}}(t)/N(t)$ and $I_{\mathrm{mt}}(t)/N(t)$ correspond to the fraction of individuals in the UK that are infected with wild-type and mutant respectively. The expectation is taken over the filtered trajectories that were reconstructed with the SMC algorithm. Finally, the scaling parameter $\lambda_0$ represents the product of the contact rate between individuals in the two countries, and the probability of infection per contact. When we fit the model to the Dutch data, the parameter $\lambda_0$ is estimated, but the fraction of infectious UK citizens is assumed known.

***Initial condition***

To complete the description of the dynamic model, we have to specify the initial conditions. Let $\zeta$ and $\xi$ denote the fractions of infected and removed individuals at time $t_0$, respectively. To determine the correct balance between exposed, infectious, and severely infected individuals, we compute the eigenvalues of the Jacobian matrix of the infinite population limit of the MJP model. The linearized system without the S and R compartments around the disease-free steady state with $S = (1-\xi)N$ and $R = \xi N$ equals

$$\frac{\mathrm{d}}{\mathrm{d}t}\begin{pmatrix} E \\ I \\ H \end{pmatrix} = \begin{pmatrix} -\alpha & \beta(1-\xi) & 0 \\ \alpha & -\gamma-\nu & 0 \\ 0 & \nu & -\omega \end{pmatrix}\begin{pmatrix} E \\ I \\ H \end{pmatrix} \tag{B-3}$$

We use the eigenvector $X_0 = (E_0, I_0, H_0)^T$ with $\sum_{i=1}^{3} X_0^i = \zeta N$ corresponding to the dominant eigenvalue of the Jacobian matrix in Eq. (B-3) to define the initial condition of the model. The parameter $p_0$ determines the initial fraction of infections with the mutant virus, and hence we have $E_{\mathrm{mt},0} = p_0 E_0$, $I_{\mathrm{mt},0} = p_0 I_0$, $E_{\mathrm{wt},0} = (1-p_0)E_0$, and $I_{\mathrm{wt},0} = (1-p_0)I_0$. We then set $S_0 = (1-\xi-\zeta)N$ and $R_0 = \xi N$. Finally, the initial state of the stochastic model is randomized by sampling from a Poisson distribution with mean equal to the deterministic initial value.

***Hybrid model simulation***

Because for small population sizes the diffusion approximation defined by the system of SDEs in Eq. (9) breaks down, as it e.g., does not allow for fixation of the mutant, we model small population sizes discretely using an adaptive tau-leap approximation of the Markov jump process (Eq. (5)). Hence, we implemented a hybrid algorithm in which variables can switch between a discrete and continuous type.

At any particular time $t$, the system consists of continuous components $X^i$ for $i \in \mathscr{C}(t)$ and discrete elements $X^i$ for $i \in \mathscr{D}(t) = \{1,\dots,n\} \setminus \mathscr{C}(t)$. Since for $i \in \mathscr{C}(t)$, the continuous element $X^i(t)$ is generally non-constant, the transition rates $\eta_j(X,t)$ will in general be time dependent. This means that we have to integrate the following system of SDEs and ODEs

$$
\begin{aligned}
\mathrm{d}X^i &= \sum_{j=1}^{k} \varepsilon_j^i \eta_j(X,t)\mathrm{d}t + \sum_{j=1}^{k} \varepsilon_j^i \sqrt{\eta_j(X,t)}\mathrm{d}B_t^j + \tau X^i \mathrm{d}\widetilde{B}_t^i\,, \quad i \in \mathscr{C}(t) \\
\frac{\mathrm{d}H^j}{\mathrm{d}t} &= \eta_j(X,t)\,, \quad j \in \mathscr{E}(t) \equiv \{j \in \{1,\dots,k\} : \exists i \in \mathscr{D}(t) : \varepsilon_j^i \neq 0\}
\end{aligned}
\tag{B-4}
$$

Hence, we have to keep track of those transition rates $\eta_j$ for which the increment $\varepsilon_j^i$ is non-zero for a discrete component $X^i$. The initial conditions for the hybrid system Eq B-4 are given by $X^i(t_m) = x_m^i$ and $H_j(t_m) = 0$. We then integrate the system until time $t_{m+1} = t_m + h_m$. At this point, we sample the number of stochastic events $Y_m$ that occurred in the time interval $(t_m, t_{m+1}]$, from the Poisson distribution

$$
Y_m \sim \text{Poisson}\left(\sum_{j \in \mathscr{E}(t_m)} H_j(t_{m+1})\right) \tag{B-5}
$$

Thereafter, $Y_m$ events with index $j$ are sampled from the categorical distribution, with probability proportional to the cumulative transition rate $H_j(t_{m+1})$. The increments $\varepsilon_j$ are then added to the discrete part of the state $X$

$$
X^j(t_{m+1}) \mapsto X^j(t_{m+1}) + \varepsilon_i^j\,, \quad i \in \mathscr{D}(t_m) \tag{B-6}
$$

After applying these $Y_m$ discrete transitions, we have to re-evaluate which components of the state are discrete and which are continuous. We choose a fixed threshold $T = 50$ below and above the populations are discrete and continuous, respectively. Hence, at time $t_{m+1}$ we update the partition of $\{1,\dots,n\}$ as follows

$$
\mathscr{D}(t_{m+1}) = \{i : X^i < T\}\,, \quad \mathscr{C}(t_{m+1}) = \{i : X^i \geq T\} \tag{B-7}
$$

Finally, we set the next initial condition $x_{m+1} = X(t_{m+1})$ and $H(t_{m+1}) = 0$ and repeat the process.

The tau-leap step size $h_m$ is chosen adaptively such that the expected number of events $\mathbb{E}[Y_m]$ within each $\tau$-leap interval $(t_m, t_{m+1}]$ is approximately equal to 1. To accomplish this, we choose

$$
h_m = \min\left\{h_{\max}, \left(\sum_{j \in \mathscr{E}(t_m)} \eta_j(X(t_m), t_m)\right)^{-1}\right\} \tag{B-8}
$$

where $h_{\max} = 1$d. Between jumps, the hybrid system (Eq B-4) is integrated using the Euler-Mayurama method with a step size of $\min\{0.01, h_m\}$.

***Sequential Monte-Carlo***

The method used for inference is described in full detail and generality elsewhere [3]. Here we give a brief description highlighting some of the choices made for this particular model and data set.

In order to reconstruct the latent epidemic trajectories $X$, given the observed data $O$, consisting of death incidence data $D$ and genetic data $F^{\mathrm{mt}}$ and $F$, we use sequential Monte-Carlo (SMC). We simulate $J = 10^4$ replicates of the model (particles) forward in time from one observation time ($t_{i-1}$) to the next ($t_i$), each with different initial conditions $X_j(t_{i-1})$. Given each of the $J$ predicted states $X_j(t_i)$ of the model at time $t_i$, we calculate the likelihood $w_j = L(O_i|X_j(t_i),\theta)$. We then sample with replacement $J$ particles with probability proportional to the weight $w_j$ using a systematic resampling method [4]. The re-sampled particles are used as initial condition at time $t_i$, and we repeat the above steps until we reach the final observation.

The Monte-Carlo estimate of the conditional likelihood of observation $O_i|O_{i-1}$, is given by the average of the weights

$$L(O_i|O_{i-1},\theta) = \frac{1}{J}\sum_{j=1}^{J} L(O_i|X_j(t_i),\theta) \tag{B-9}$$

where we write $L(O_1|O_0,\theta) \equiv L(O_1|\theta)$. The total likelihood of the time series given $\theta$ is equal to

$$L(O_1,\ldots,O_N|\theta) = \prod_{i=1}^{N} L(O_i|O_{i-1},\theta) \tag{B-10}$$

All likelihood computations are done on the log-scale to minimize floating-point errors.

To estimate parameters, we extended the state $X$ with the parameter vector $\theta$, allowing the parameters to be perturbed after each observation time. For the $j$-th particle, we now have a state $(X_j,\theta_j)$, and weight $w_j = L(O_i|X_j(t_i),\theta_j)$. The extended-SMC algorithm is then iterated $M = 200$ times, and after each iteration $m$ the magnitude of the parameter perturbations is reduced. The perturbations are Gaussian $\theta_j \mapsto \theta_j + a^m\varepsilon$, with $\varepsilon \sim \mathcal{N}(0,\Sigma)$. Here, $\Sigma$ is a diagonal matrix with appropriately chosen diagonal elements. For bounded parameters, the perturbations are reflected in the boundary of the domain. The ratio $a \in (0,1)$ reduces the magnitude of the parameter perturbations. We choose $a = \sqrt[M]{10^{-2}}$ such that after $M$ iterations the magnitude of the perturbations is reduced by 99%. After each iteration of the extended-SMC algorithm, for each particle we reset the state $X_j$ to the a randomly sampled initial state of the epidemic model, while $\theta_j$ is inherited from the previous extended-SMC iteration. To speed-up the computations, we implemented the model and SMC algorithm in C++ and used multi-threading to update particles between observations in parallel.

#### Appendix C: The effect of NPI on the estimate of $s$

In both our population genetics and epidemic model, we estimate a parameter $s$ that we interpret as the selective advantage of a new variant over background. As the two models are quite different, the meaning and estimated value of $s$ in the context of the two models is also different.

If we ignore migration, the population-genetics model is essentially a regression model with a logistic link function and a binomial likelihood

$$F_n^{\mathrm{mt}} \sim \mathrm{Binom}(F_n, p_{t_n}) \quad p_t = (1+\exp(-at-b))^{-1} \tag{C-1}$$

If both variants are growing exponentially with rate $r_{\mathrm{wt}}$ and $r_{\mathrm{mt}}$, the regression coefficient $a$ is equal to the difference in exponential growth rates, as

$$p_t = \frac{p_0 \exp(r_{\mathrm{mt}} t)}{p_0 \exp(r_{\mathrm{mt}} t) + (1-p_0)\exp(r_{\mathrm{wt}} t)} = \frac{1}{1+\exp(-(r_{\mathrm{mt}} - r_{\mathrm{wt}})t - \mathrm{logit}(p_0))} \tag{C-2}$$

and hence, $a = r_{\mathrm{mt}} - r_{\mathrm{wt}}$ and $b = \mathrm{logit}(p_0)$.

Our epidemic model is based on the SEIR model, in which the growth rate is the largest root of the quadratic polynomial [5]

$$x \mapsto (x+\alpha)(x+\gamma) - \alpha\tilde{\beta}S \tag{C-3}$$

where $\tilde{\beta} = \beta$ for the wild type, and $\tilde{\beta} = \beta(1+s)$ for the mutant. Therefore, we find that

$$a = \frac{1}{2}\left(\sqrt{(\alpha-\gamma)^2 + 4\alpha\beta(1+s)S} - \sqrt{(\alpha-\gamma)^2 + 4\alpha\beta S}\right) \tag{C-4}$$

For small $s$, we can use a first-order Taylor expansion to derive

$$\begin{aligned} a &= \frac{\alpha\beta S s}{\sqrt{(\alpha-\gamma)^2 + 4\alpha(\beta S - \gamma)}} + \mathcal{O}(s^2) \\ &= s\frac{R_e}{T_G}\frac{1}{\sqrt{1+4\frac{R_e - 1}{T_G(\alpha+\gamma)}}} + \mathcal{O}(s^2) \end{aligned} \tag{C-5}$$

where the generation time $T_G$ is given by $T_G = 1/\alpha + 1/\gamma$ [6] and $R_e = \beta S/\gamma$ is the effective reproduction number of the wild-type. Therefore, when $R_e$ is close to 1, we have $aT_G \approx s$, meaning that the selective advantage per generation in the two models are comparable. However, when $R_e > 1$ the population genetics model estimates a larger selective advantage that the epidemic model.

Non-pharmaceutical interventions (NPI) reduce the effective contact rate between individuals, which is implemented as a reduction in the parameter $\beta$ in the epidemic model. Even though these NPI act equally on both variants (by assumption), we will show here that effective NPI can influence the value of $s$ that we estimate with the population genetics model. This can explain some of the variation in $s$ observed between countries, as governments have implemented NPI differently.

Consider the following deterministic simplification of the epidemic model

$$\begin{aligned} \frac{\mathrm{d}S}{\mathrm{d}t} &= -\beta S(I_{\mathrm{wt}} + (1+s)I_{\mathrm{mt}}) \\ \frac{\mathrm{d}E_{\mathrm{wt}}}{\mathrm{d}t} &= \beta S I_{\mathrm{wt}} - \alpha E_{\mathrm{wt}} \\ \frac{\mathrm{d}E_{\mathrm{mt}}}{\mathrm{d}t} &= \beta(1+s)S I_{\mathrm{mt}} - \alpha E_{\mathrm{mt}} \\ \frac{\mathrm{d}I_{\mathrm{wt}}}{\mathrm{d}t} &= \alpha E_{\mathrm{wt}} - \gamma I_{\mathrm{wt}} \\ \frac{\mathrm{d}I_{\mathrm{mt}}}{\mathrm{d}t} &= \alpha E_{\mathrm{mt}} - \gamma I_{\mathrm{mt}} \end{aligned} \tag{C-6}$$

with initial condition $S(0) = 1-\zeta$, and a fraction $\zeta$ individuals is distributed over the remaining classes using the dominant eigenvector of the Jacobian of the Eq. (C-6) and initial mutant frequency $p_0$. The parameter $\beta$ is a function of time, given by $\beta(t) = (1 - H_\upsilon(t-t_1))\beta_0 + H_\upsilon(t-t_1)\beta_1$ where $H_\upsilon(t) = (1+e^{-t/\upsilon})^{-1}$ is a smoothed step function. Hence, before NPI are implemented, the infection rate is $\beta_0$, and $\beta_1$ afterwards. By simulating this model, we can generate synthetic data $(F_n^{\mathrm{wt}}, F_n^{\mathrm{mt}})$ at observation time $t_n$ sampling as follows

$$F_n \sim \mathrm{Poisson}(M)\,, \quad F_n^{\mathrm{mt}} \sim \mathrm{Binom}(F_n, f_{\mathrm{mt}}(t_n))\,, \quad \text{and} \quad F_n^{\mathrm{wt}} = F_n - F_n^{\mathrm{mt}} \tag{C-7}$$

The expected frequency $f_{\mathrm{mt}}$ of sampled mutant sequences is approximated with

$$f_{\mathrm{mt}}(t) = \frac{E_{\mathrm{mt}}(t)}{E_{\mathrm{mt}}(t) + E_{\mathrm{wt}}(t)} \tag{C-8}$$

We simulated the simplified model for different values of the effectiveness of the NPI $\rho \equiv \beta_1/\beta_0$. The trajectories of the faction of infectious individuals is shown in Fig. S1A, and the mutant frequency $f_{\mathrm{mt}}$ is shown in Fig. S1B. This shows that effective NPI has a diminishing effect on the increase of the mutant frequency. We then used the population genetics model to estimate the selective advantage $s$ for the different values of $\rho$. An example fit is shown in Fig. S1C and the posterior densities of $s$ are shown in Fig. S1D.

#### Literature Cited


[1] Mumford, D., Series, C. & Wright, D. *Indra's Pearls: The Vision of Felix Klein*. Cambridge University Press (2002).

[2] Rozhnova, G. et al. Model-based evaluation of school- and non-school-related measures to control the COVID-19 pandemic. *Nature Communications* **12**, 1614 (2021).

[3] Ionides, E. L., Nguyen, D., Atchadé, Y., Stoev, S. & King, A. A. Inference for dynamic and latent variable models via iterated, perturbed Bayes maps. *Proceedings of the National Academy of Sciences* **112**, 719–724 (2015).

[4] Douc, R. & Cappe, O. Comparison of resampling schemes for particle filtering. In *ISPA 2005. Proceedings of the 4th International Symposium on Image and Signal Processing and Analysis, 2005.*, 64–69 (2005).

[5] Ma, J. Estimating epidemic exponential growth rate and basic reproduction number. *Infect Dis Model* **5**, 129–141 (2020).

[6] Roberts, M. G. & Heesterbeek, J. A. Model-consistent estimation of the basic reproduction number from the incidence of an emerging infection. *J Math Biol* **55**, 803–816 (2007).

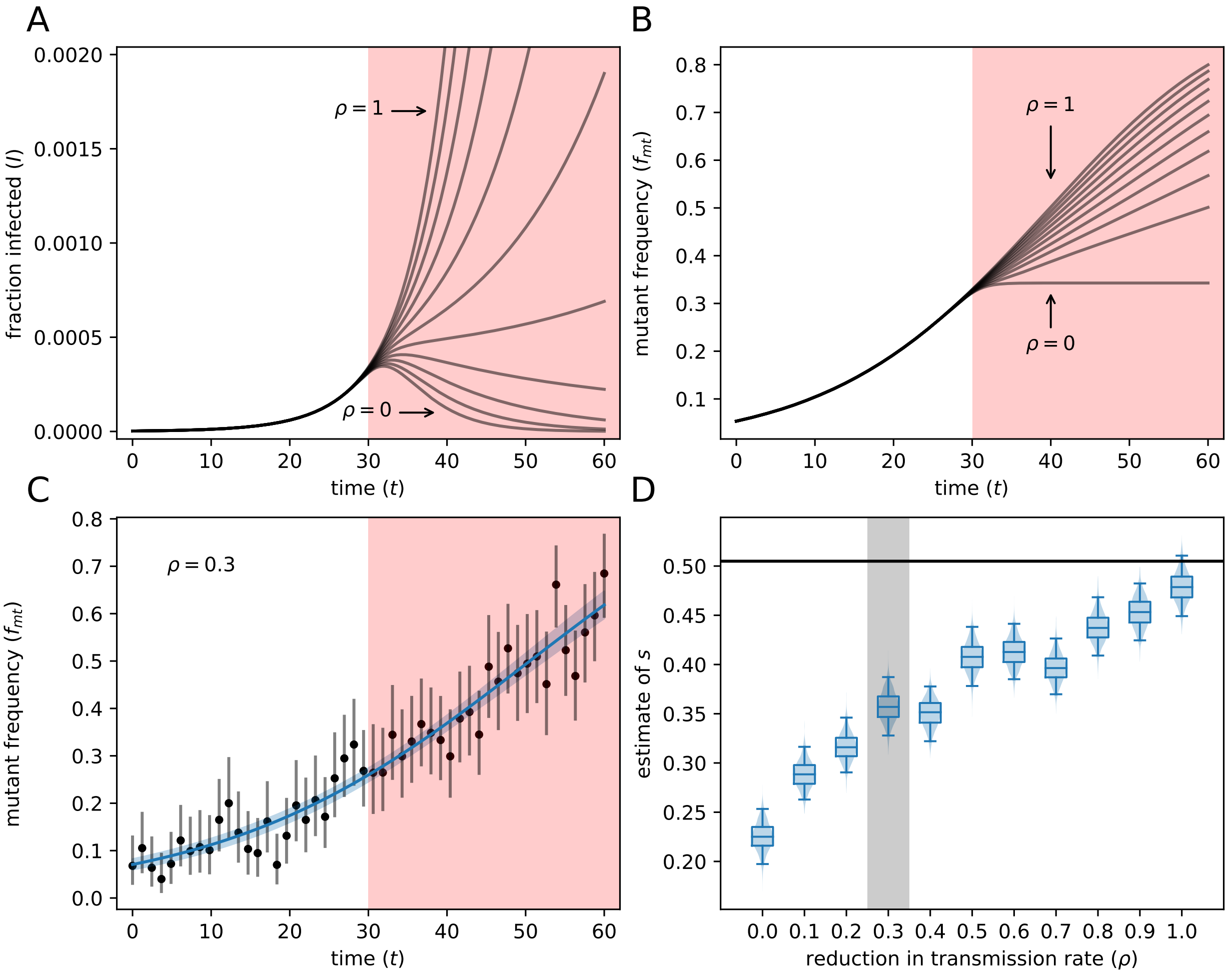


Figure S1: **Effective NPI negatively biases the value of $s$ estimated with the population genetics model.** Panel A shows the trajectory of the number of infectious individuals for 10 different values of $\rho \equiv \beta_1/\beta_0$, the ratio of transmission rates after and before NPI. The pink area represents implementation of NPI. Panel B shows the fraction of infections with the mutant. Panel C shows simulated data (black dots and gray 95% CIs) and the fit of the model (mean and 95% CrI in blue) in the case $\rho = 0.3$. Panel D shows the estimated value of $s$ for the various values of $\rho$. The black line shows the ground-truth value of $s$, corrected with Eq. (C-4). The gray area indicates the estimate from the data shown in panel C. Parameters: $\beta_0 = 0.6$, $t_1 = 30$, $\zeta = 5 \cdot 10^{-6}$, $\alpha = 1/3$, $\gamma = 1/4$, $s = 0.35$, $p_0 = 0.05$, $M = 100$.

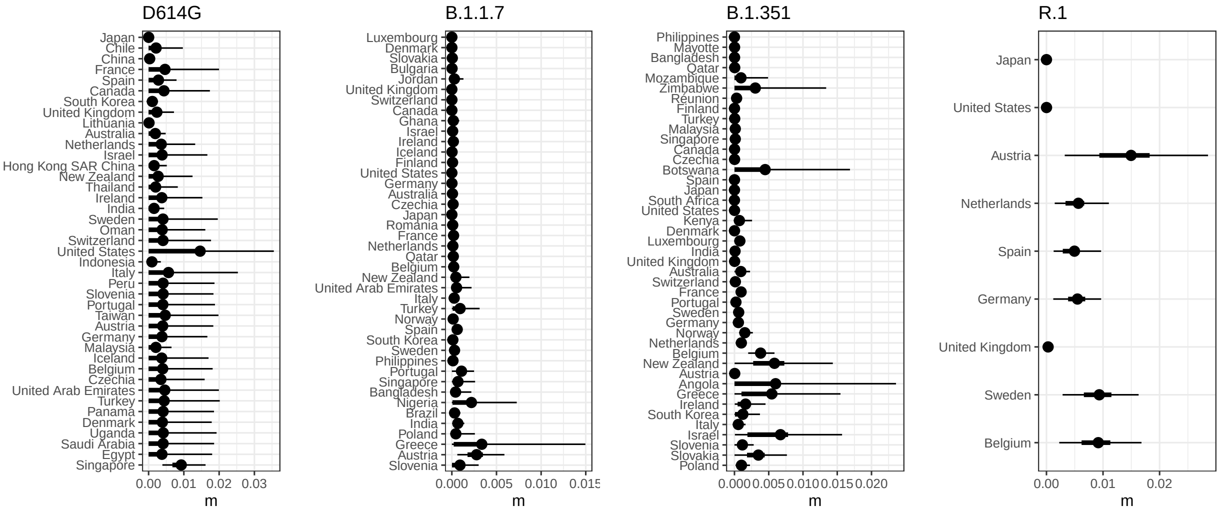


Figure S2: Migration fractions for each country from the population genetic model. Results are for the final time points shown in Figs. 2–4. Points mark the median, and thick and thin lines are 50% and 95% CIs, respectively. Corresponding estimates of migration are in Fig. S2.

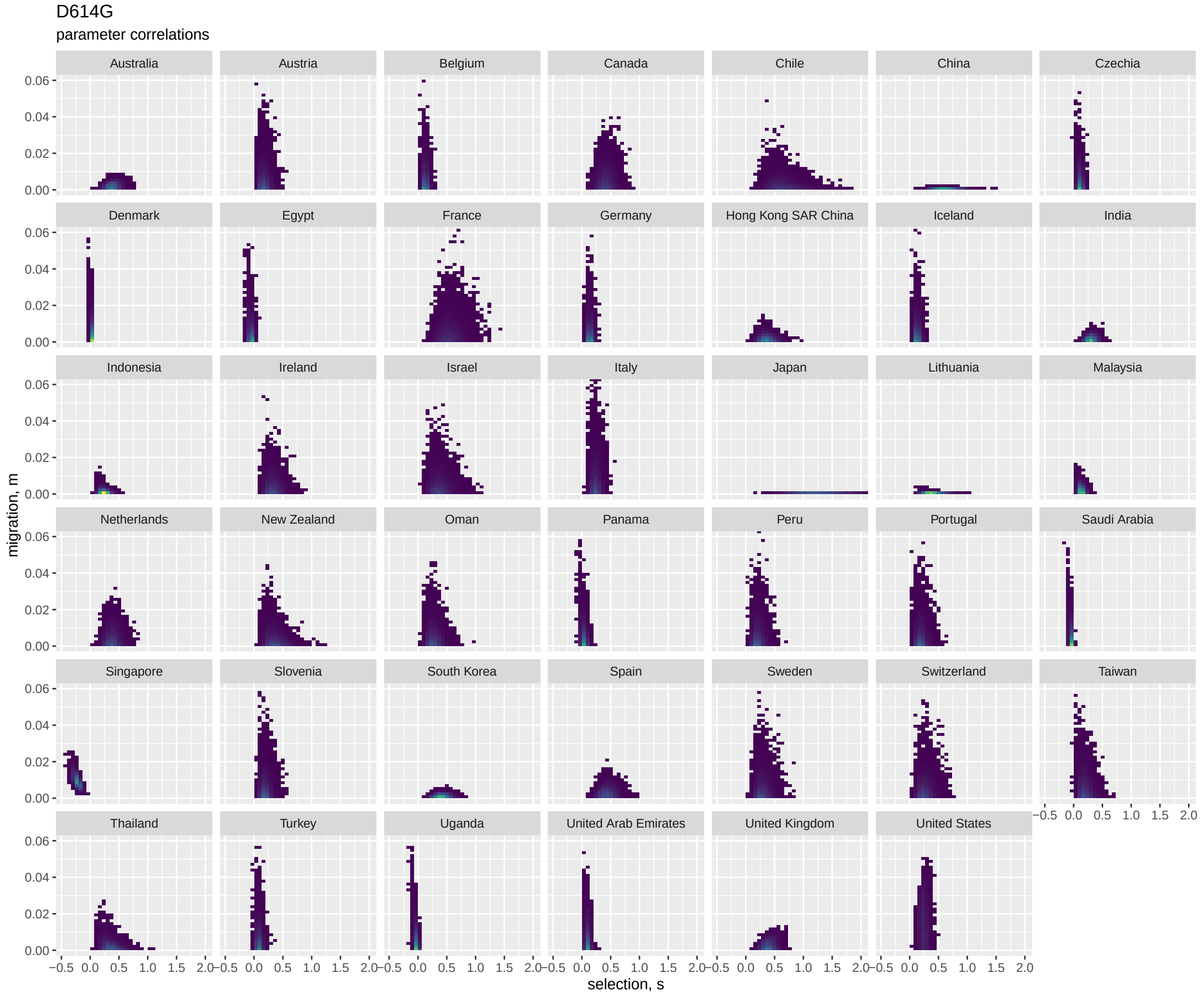


Figure S3: Correlations between the selection and migration parameters from the population genetic model, for D614G at the final time point. The scatter plots represent samples from the joint posterior distribution of $s$ and $m$.

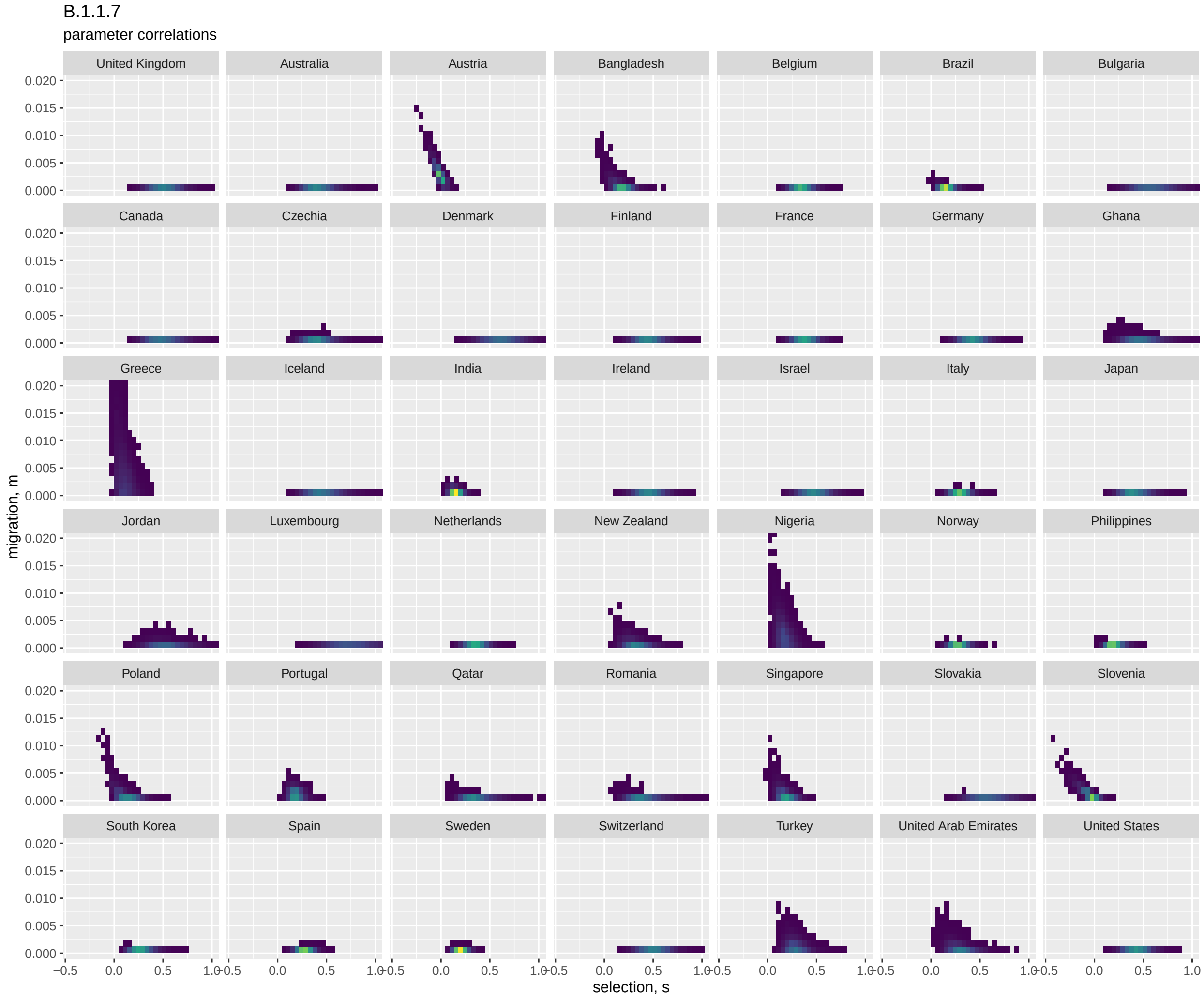


Figure S4: Correlations between the selection and migration parameters from the population genetic model, for B.1.1.7 at the final time point. The scatter plots represent samples from the joint posterior distribution of $s$ and $m$.

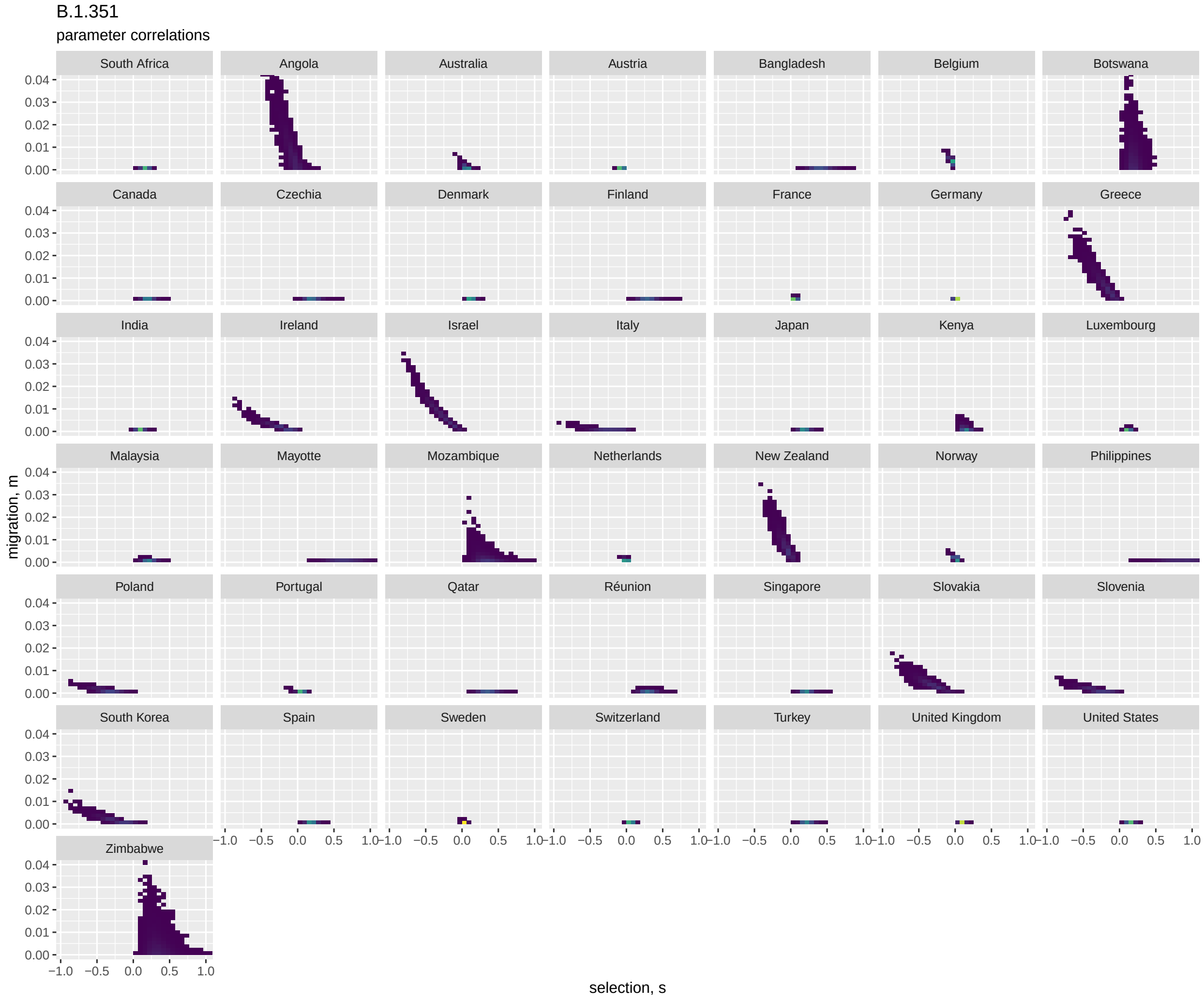


Figure S5: Correlations between the selection and migration parameters from the population genetic model, for B.1.351 at the final time point. The scatter plots represent samples from the joint posterior distribution of $s$ and $m$.

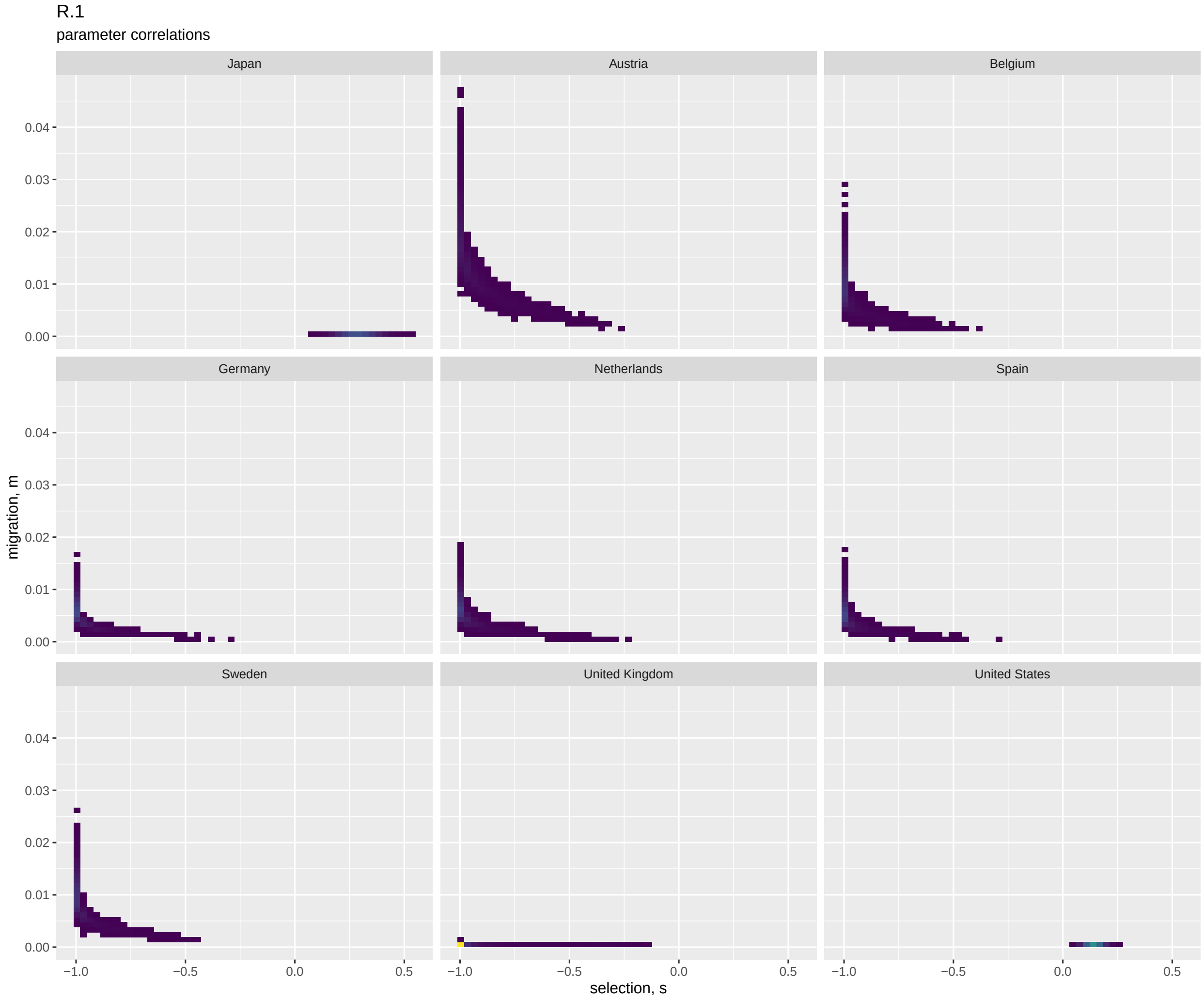


Figure S6: Correlations between the selection and migration parameters from the population genetic model, for R.1 at the final time point. The scatter plots represent samples from the joint posterior distribution of $s$ and $m$.

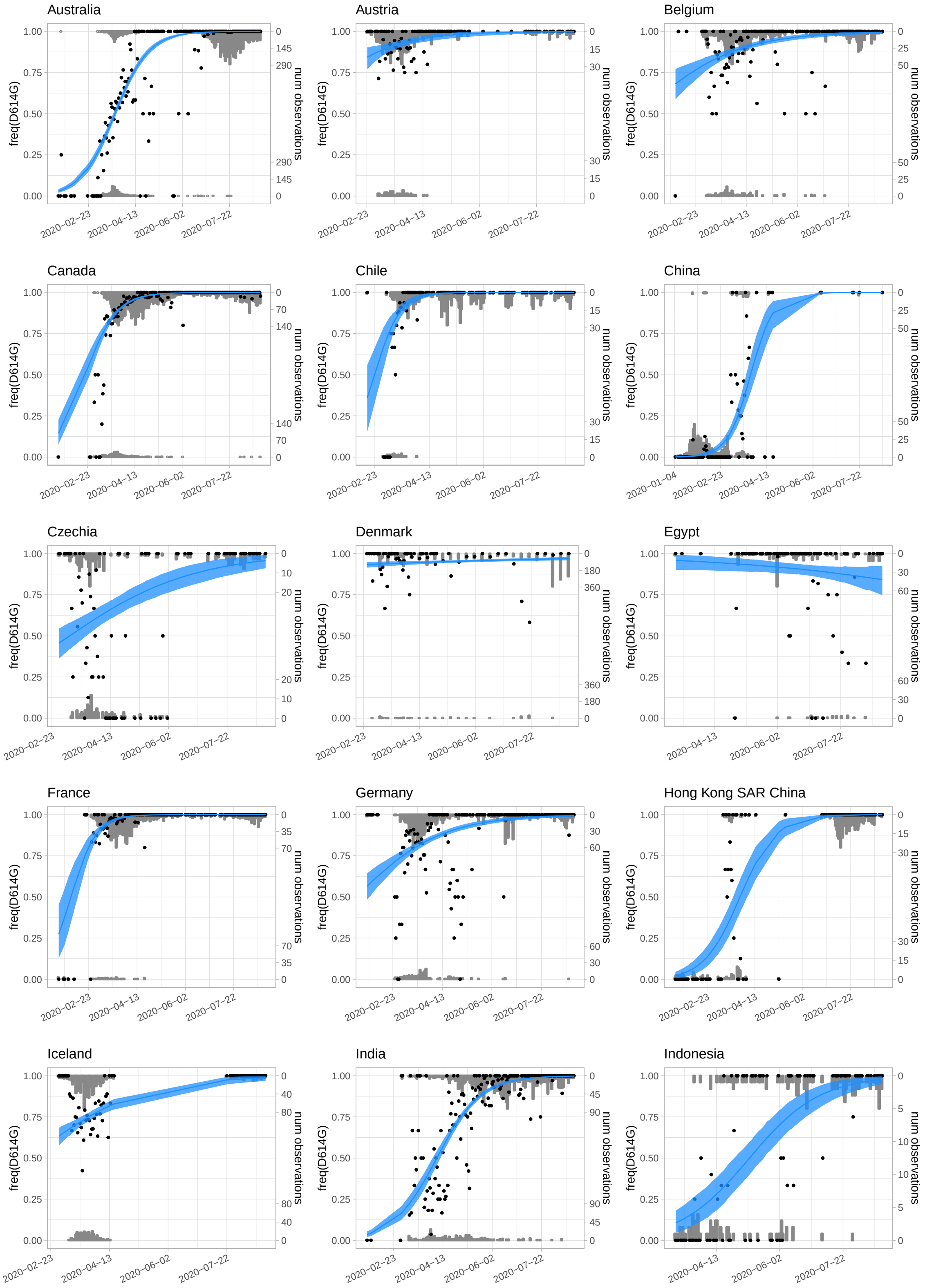


Figure S7: Fits from the hierarchical population genetic model to the daily variant data for D614G, up to the final point. Gray bars show histograms of numbers of observations of focal variant (upper axis) and all other variants (lower axis) each day. Black dots are the daily frequencies. Blue shows the frequencies predicted from the model, with 95% credible intervals shaded.

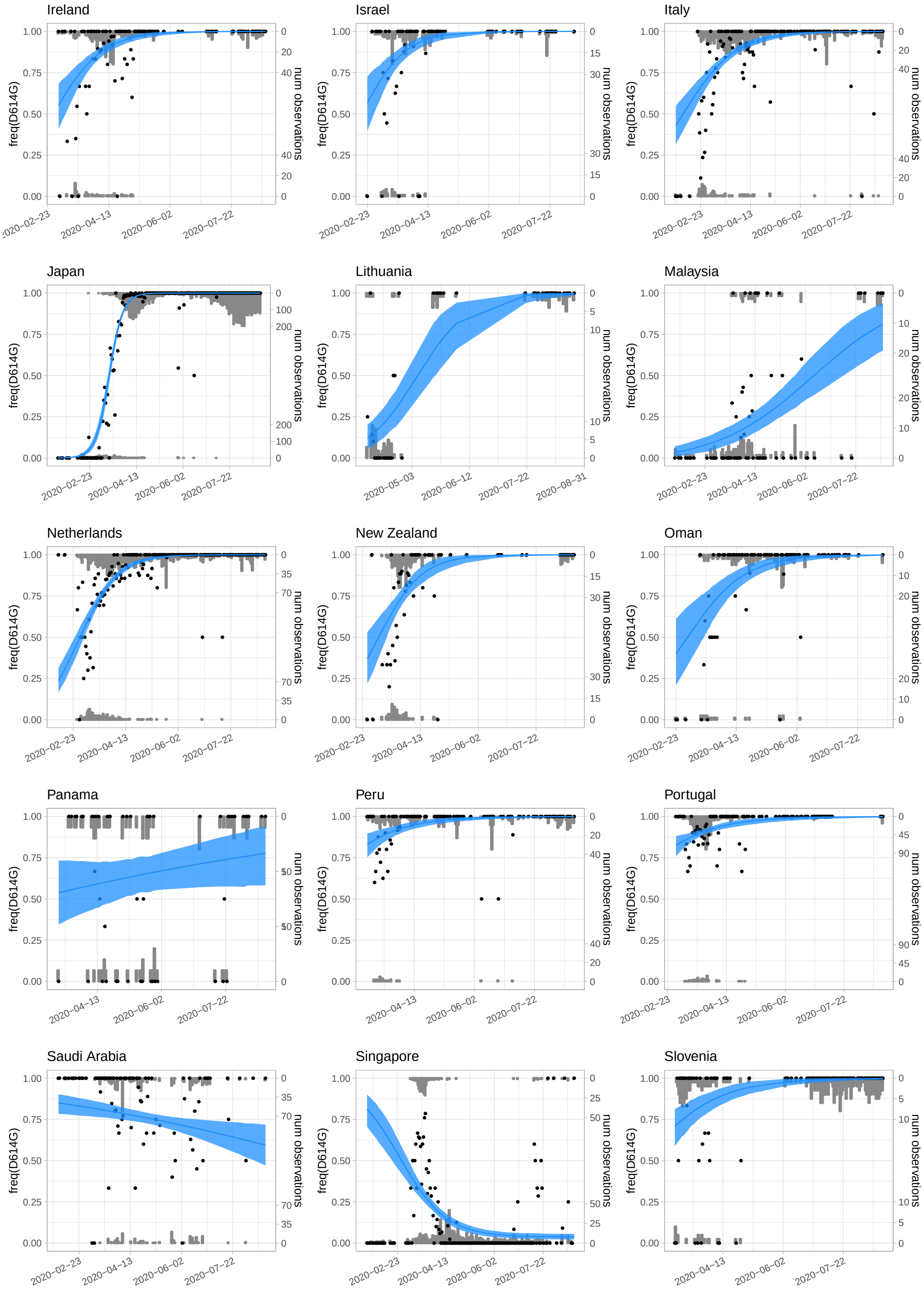


Figure S7: (continued)

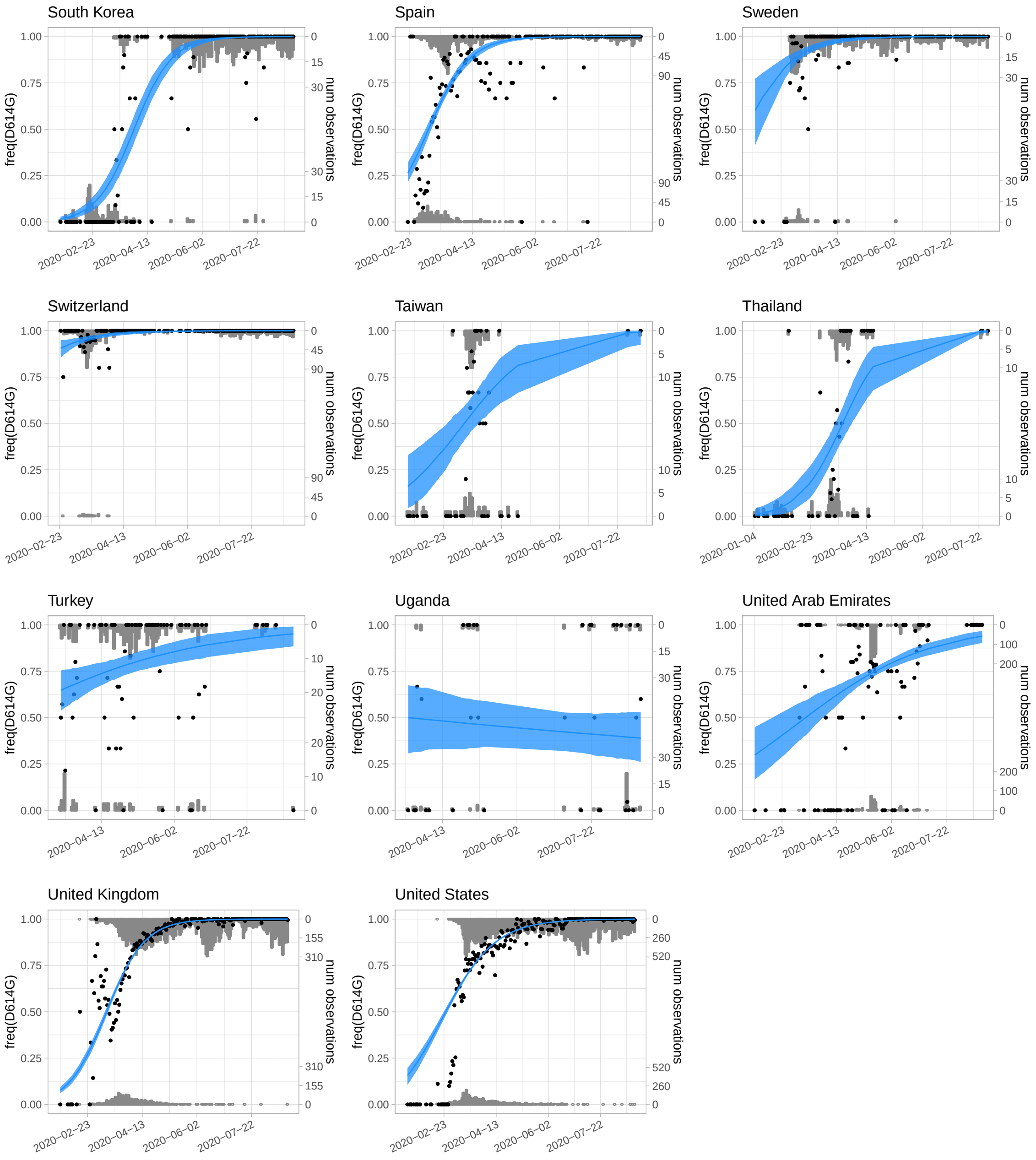


Figure S7: (continued)

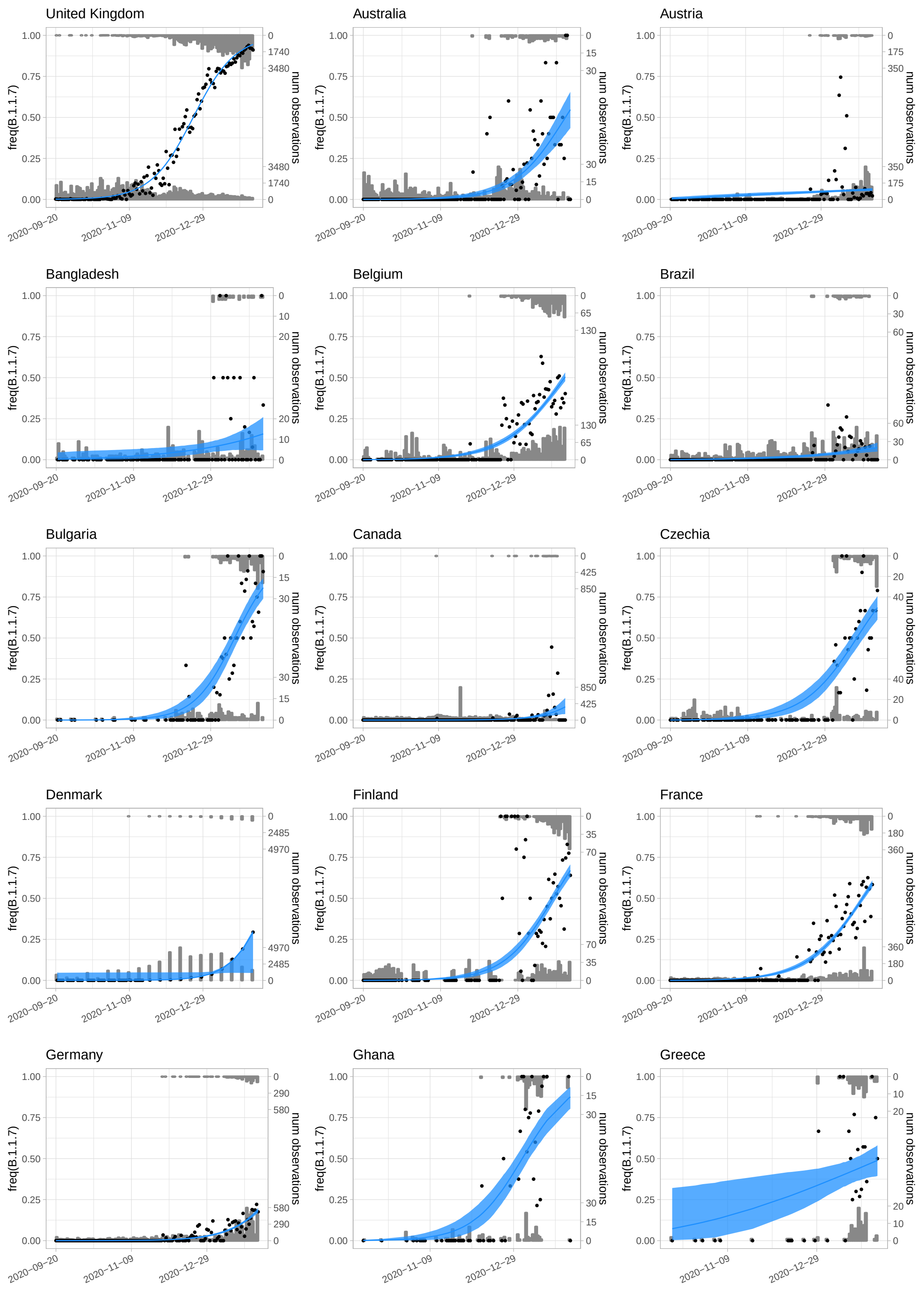


Figure S8: Fits from the hierarchical population genetic model to the daily variant data for B.1.1.7, up to the final time point. Gray bars show histograms of numbers of observations of B.1.1.7 (upper axis) and all other variants (lower axis) each day. Black dots are the daily frequencies. Blue shows the frequencies predicted from the model, with 95% credible intervals shaded.

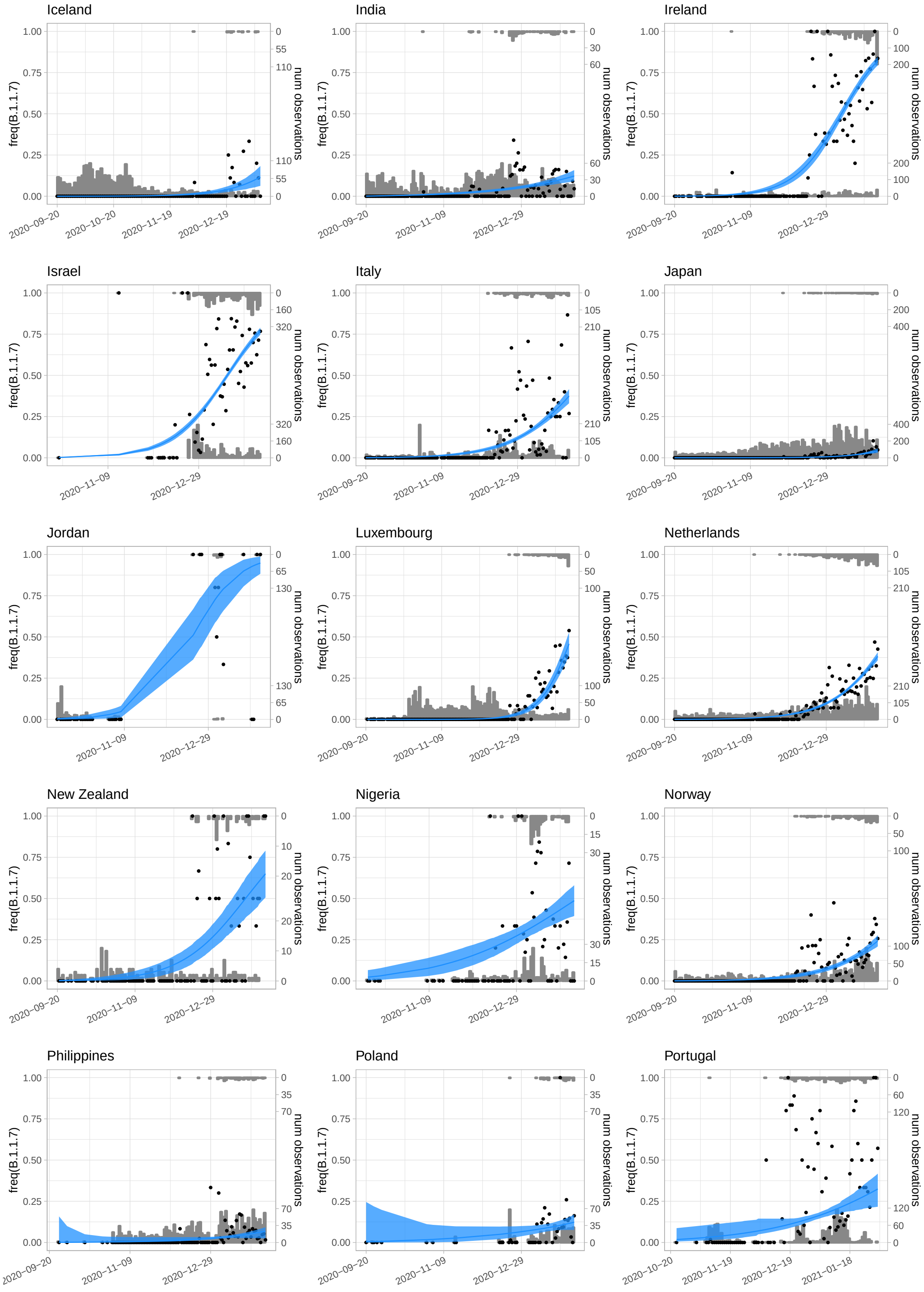


Figure S8: (continued)

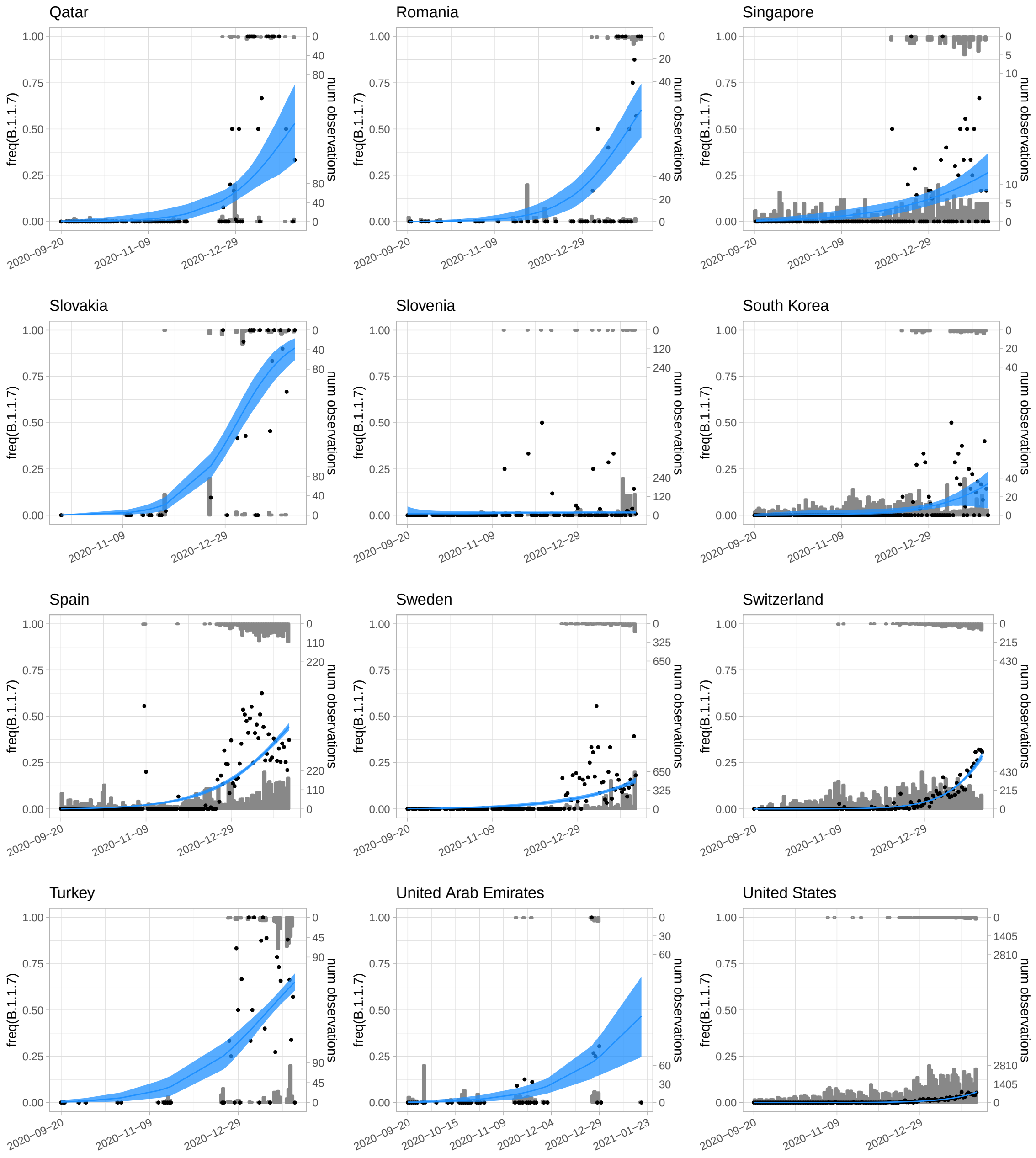


Figure S8: (continued)

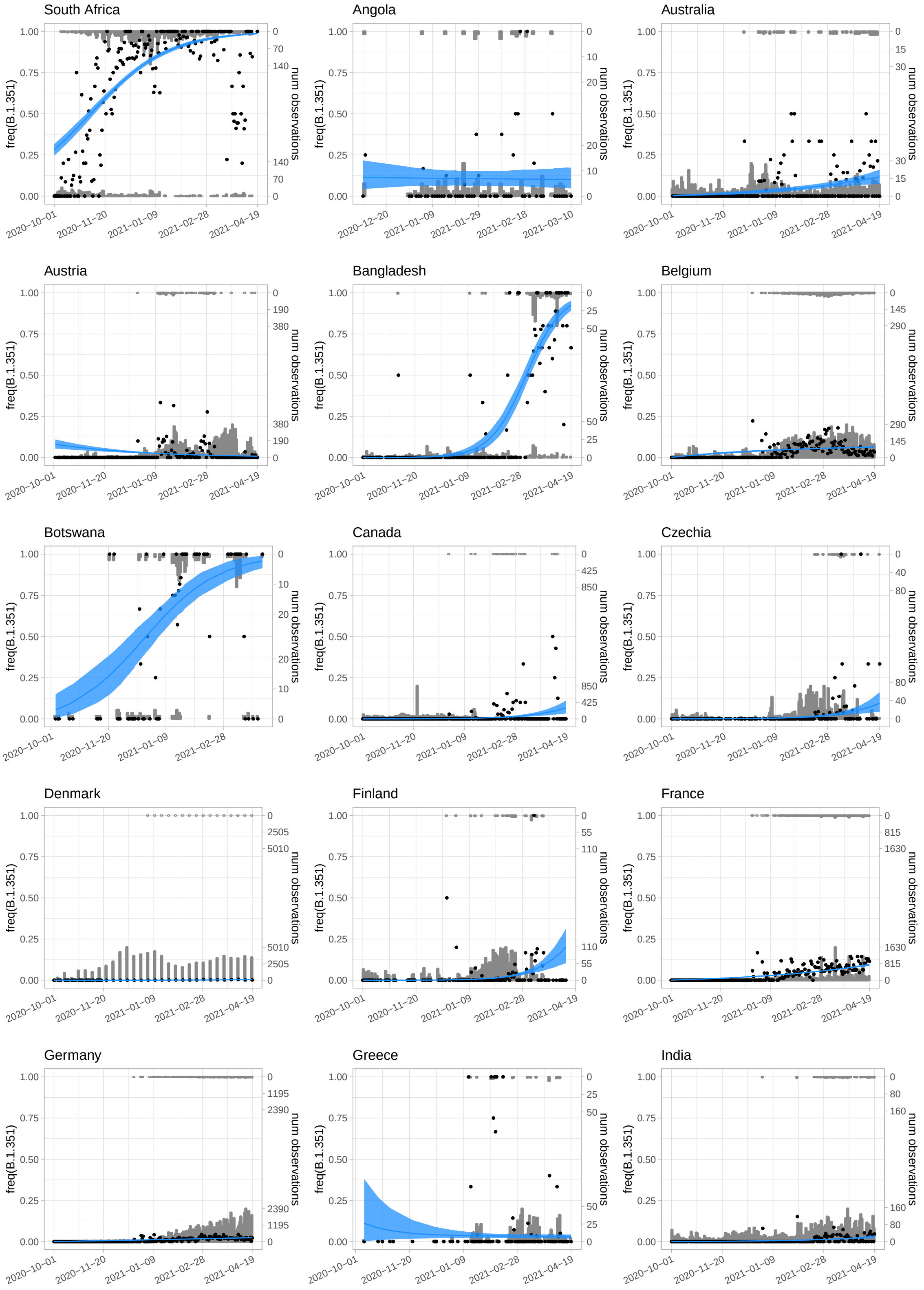


Figure S9: Fits from the hierarchical population genetic model to the daily variant data for B.1.351, up to the final time point. Gray bars show histograms of numbers of observations of the focal variant (upper axis) and all other variants (lower axis) each day. Black dots are the daily frequencies. Blue shows the frequencies predicted from the model, with 95% credible intervals shaded.

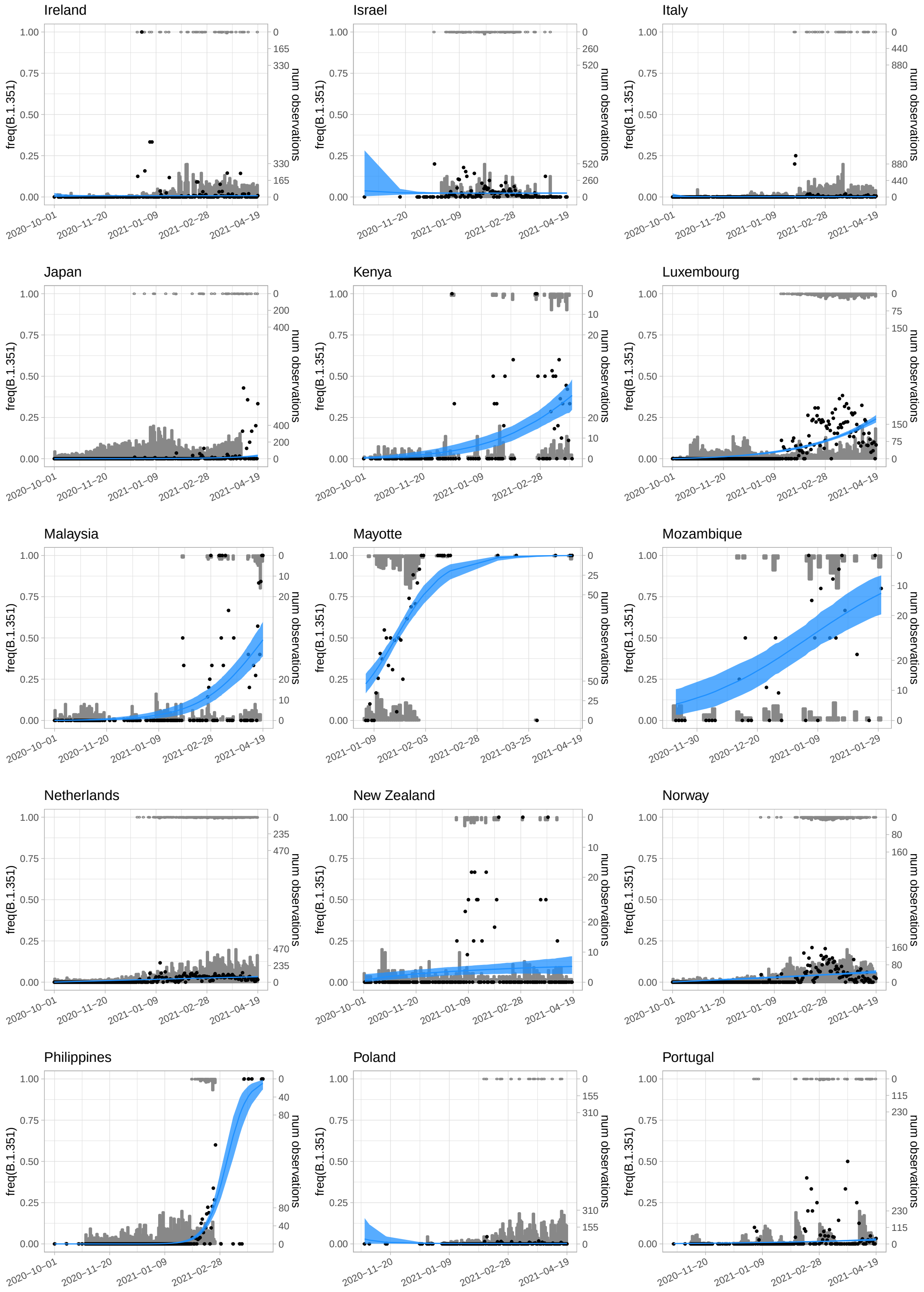


Figure S9: (continued)

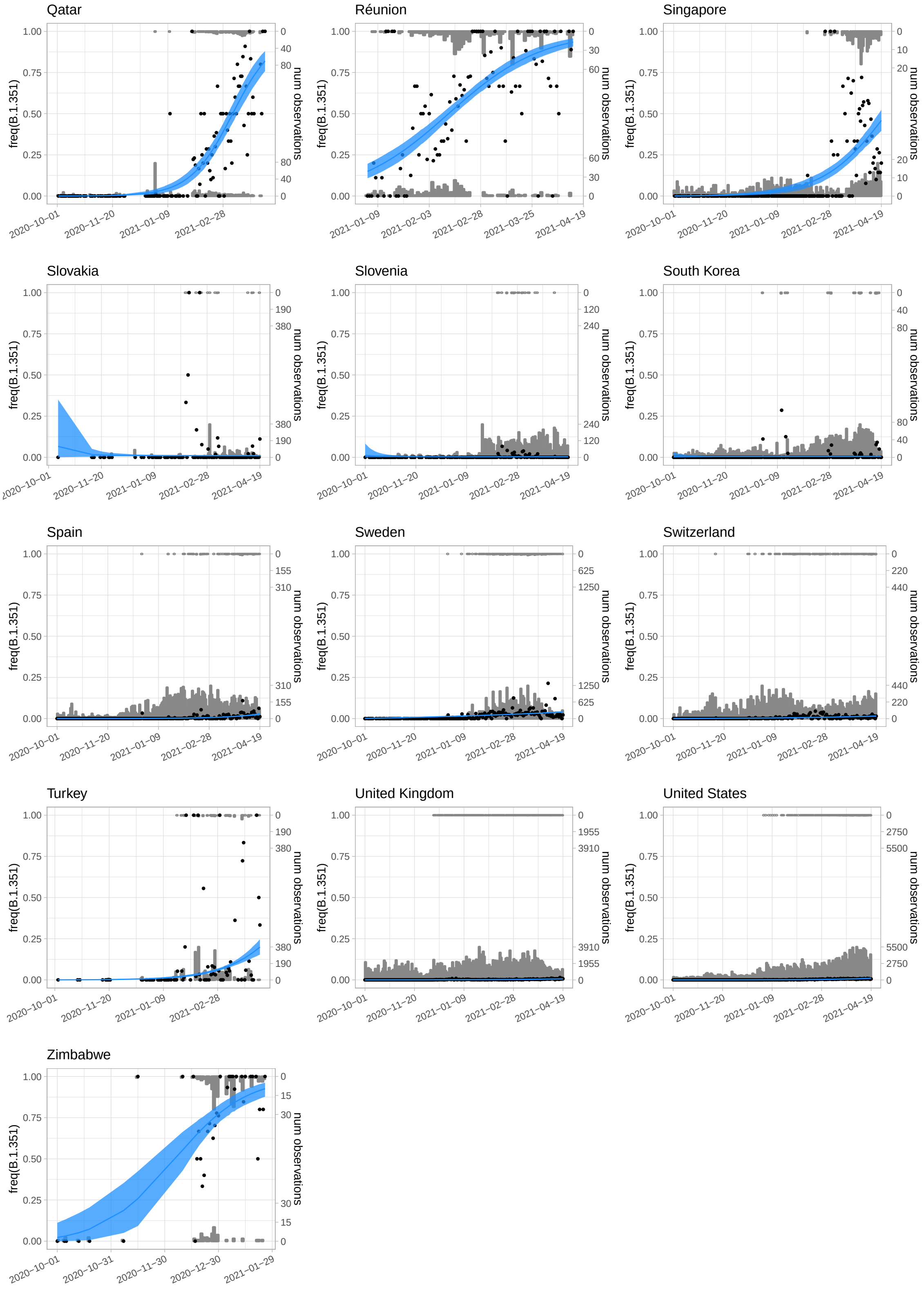


Figure S9: (continued)

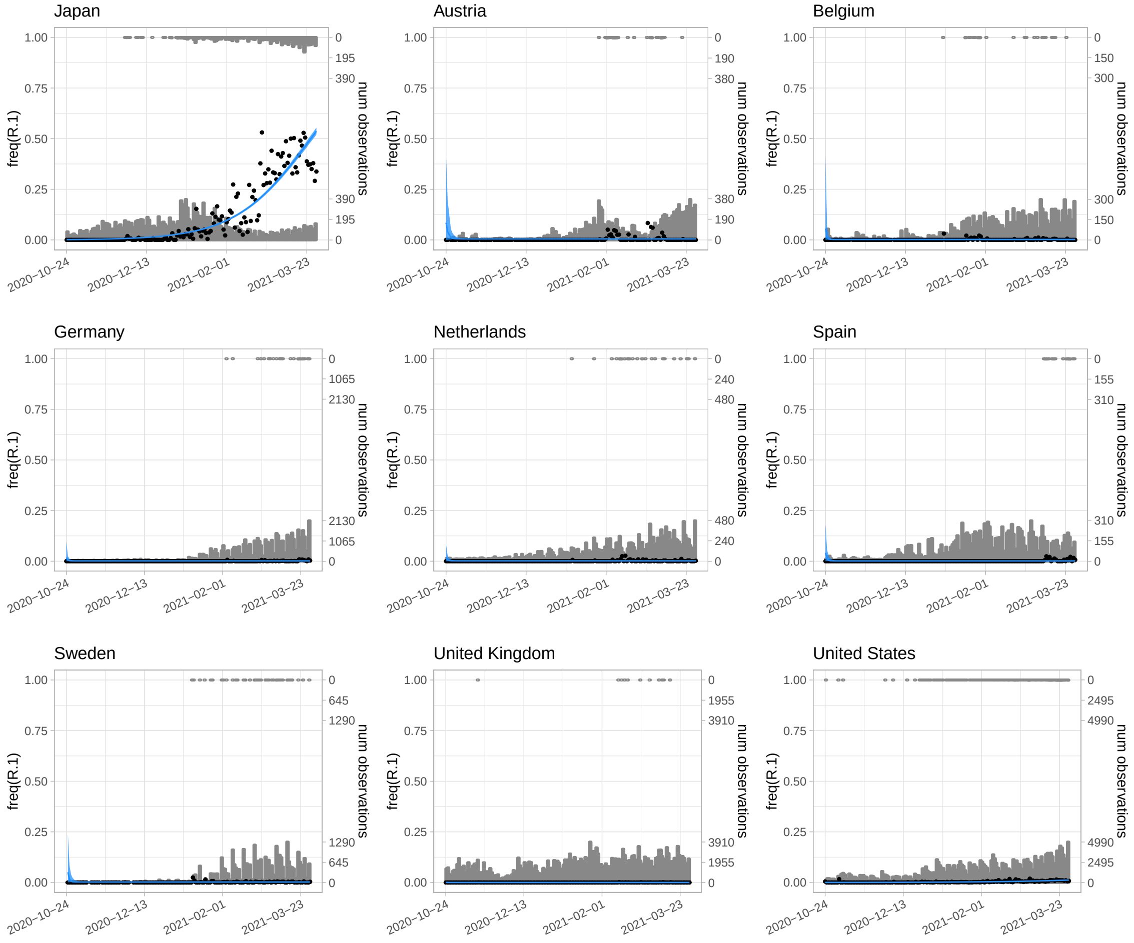


Figure S10: Fits from the hierarchical population genetic model to the daily variant data for R.1, up to the final time point. Gray bars show histograms of numbers of observations of the focal variant (upper axis) and all other variants (lower axis) each day. Black dots are the daily frequencies. Blue shows the frequencies predicted from the model, with 95% credible intervals shaded.

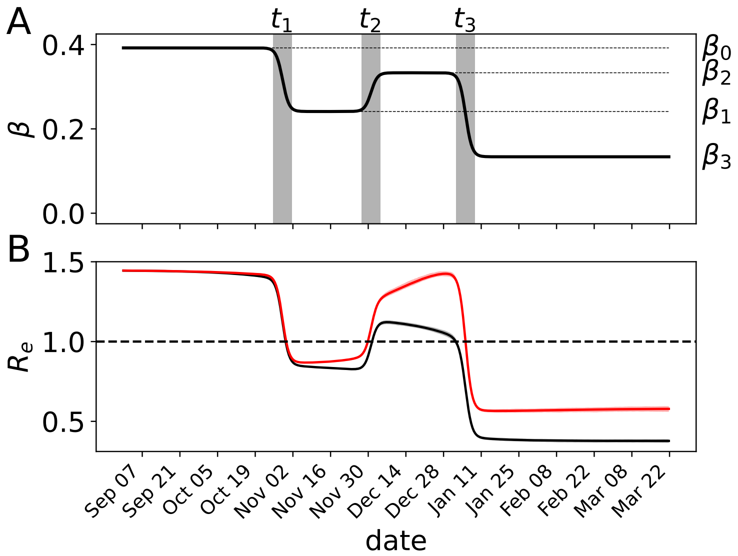


Figure S11: Inferred effective reproduction number in the UK. A. The infection rate $\beta$ is time-dependent to model government responses to the pandemic. The uptake-period of changes in restrictions is shown in gray. Parameters used for this example are form the UK and variant B.1.1.7. B. Using the reconstructed trajectories of the fitted stochastic epidemic model, the instantaneous effective reproduction number ($R_e$) is calculated as $R_e = \tilde{\beta}/(\gamma + \nu)S/N$. The lighter bands indicate the 2.5 - 97.5 percentiles of the filtered trajectories. The thick lines indicate the mean. The effective reproduction number for the wild-type is shown in black ($\tilde{\beta} = \beta$). The red curve indicates the weighted average of the effective reproduction numbers of the wild-type and mutant ($\tilde{\beta} = \beta(1 + sI_{\mathrm{mt}}/I)$). The threshold value of $R_e = 1$ secondary infections per infected individual is indicated by the horizontal, dashed line.
